# Health care resource consumption and corresponding economic burden in patients with acute cardiovascular diseases during the COVID-19 pandemic period

**DOI:** 10.64898/2025.12.31.25343267

**Authors:** P Moulaire, T Delory, A Rachas, M Espagnacq, M Khlat, S Le Cœur, G Hejblum, N Lapidus, the COVID-HOSP study group

**Affiliations:** Sorbonne Université, INSERM, Institut Pierre Louis d’Épidémiologie et de Santé Publique, Paris, France; Centre Hospitalier Annecy-Genevois, Epagny Metz-Tessy, France; French Institute for Demographic Studies (INED), Mortality, Health and Epidemiology Unit, Aubervilliers, France; Caisse Natl Assurance Malad, Dept Pathol & Patients, Paris, France; Institute for Research and Information in Health Economics (IRDES), Paris, France; Sorbonne Université, INSERM, Institut Pierre Louis d’Épidémiologie et de Santé Publique, AP-HP, Hôpital Saint-Antoine, Unité de Santé Publique, Paris, France

**Keywords:** Cardiovascular, Acute Disease, Healthcare Costs, Health Administrative Data, Economic Burden, COVID-19, Pandemic

## Abstract

**Background:** During the COVID-19 pandemic period, healthcare systems substantially reorganized their management of several diseases, including acute cardiovascular diseases (ACVD) such as heart failure, stroke, myocardial infarction, and pulmonary embolism. While previous studies have reported changes in hospitalization rates and clinical outcomes, the economic impact of the pandemic on healthcare expenditures for patients with ACVD remains poorly documented. This study aimed to quantify disruptions in healthcare utilization and reimbursement trends among patients with ACVD in France during the pandemic period (2020–2023).

**Methods:** Using comprehensive French healthcare reimbursement data from 2015 to 2023, this nationwide cohort study analyzed 3.9 million ACVD-related patient-years, totaling €86 billion in reimbursements. A two-step approach was employed: first, a linear regression model based on pre-pandemic trends (2015–2019) was used to project expected expenditures for the years 2020–2023, adjusting for age, sex, calendar year, and comorbidities. Second, expenditures observed during the years 2020-2023 were compared with these projections to estimate potential disruptions related to the pandemic period. Analyses were stratified across 21 expenditure categories.

**Results:** Between 2020 and 2023, ACVD-related healthcare expenditures exceeded expected values by €2.3 billion (+6.2%), with the largest gap in 2023 (€1.1 billion above projections). Notably, pre-pandemic annual expenditure growth (€86–212 per patient) sharply accelerated during the pandemic period (€492–1320 per patient). Excess spending was higher in males (€1.4 billion), patients with severe comorbidities (*≥*3 comorbidities: €1.4 billion), and in the 65,195 patients with ACVD and a history of a COVID-19-related hospitalization(s) (€0.9 billion, driven primarily by short stay hospitalizations and rehabilitation care). Among non-COVID-19 ACVD patients, significant increases were observed in drug expenses, short stay hospitalizations, and hospitalizations in psychiatry.

**Conclusion:** In France, the COVID-19 pandemic years were marked by substantial and sustained disruptions in healthcare expenditures among patients with ACVD, extending beyond care directly related to COVID-19. Excess costs were linked both to pandemic-related complications and broader systemic shifts, including increased psychiatric and rehabilitation needs. These findings have critical public health implications: they highlight the need to address ongoing healthcare system disruptions for this vulnerable population while also reinforcing vigilance in future health crises.

## Introduction

The emergence of COVID-19 in Wuhan, China, in late 2019[1], marked the beginning of an unprecedented worldwide health crisis. SARS-CoV-2 virus rapidly spread across the world, leading the World Health Organization (WHO) to declare it a public health emergency of international concern (PHEIC) on January 30, 2020[2]. Beyond its profound impact on morbidity and mortality, with 18.2 million deaths worldwide only between Jan 1, 2020, and Dec 31, 2021[3], the pandemic has also exerted significant pressure on healthcare systems and economies worldwide[4]. Understanding the economic implications of the pandemic is crucial, as it provides insights into the resilience and adaptability of healthcare systems during crises.

Cardiovascular diseases (CVD) already represented a substantial economic burden on healthcare systems before the pandemic[5, 6]. In France, €14 billion was attributed to the care of 4 million people with CVD in 2017[7]. Acute cardiovascular conditions, in particular, require urgent and intensive medical interventions, leading to high healthcare costs. The COVID-19 pandemic caused critical disruptions in the management of acute cardiovascular diseases (ACVD), characterized by a reduction in hospitalization rates alongside an increase in in-hospital mortality [8, 9]. In France, changes in healthcare pathways of patients hospitalized for acute heart failure[10], stroke[11], myocardial infarction[12], and pulmonary embolism[13] have also been highlighted.

Although previous studies have examined the impact of the pandemic on hospitalization and clinical outcomes, to the best of our knowledge, information on healthcare utilization and related expenditures is currently lacking. Several international studies have already investigated the the impact of the COVID-19 pandemic on healthcare costs for cardiovascular conditions. However, these analyses have primarily focused on co-infections[14] or hospitalizations for COVID-19 in patients with pre-existing cardiovascular disease[15, 16]. In France, existing research on the economic burden of the pandemic has only described the costs associated with hospitalizations for COVID-19[17, 18] or long COVID[19]. To date, no study has provided a detailed assessment of the pandemic’s economic impact on acute cardiovascular diseases specifically. Quantifying these changes is essential to assess the full impact of the pandemic on healthcare systems and to inform future preparedness strategies for this vulnerable population.

Given the observed decline in hospital admissions for these conditions in 2020, one hypothesis is that the most severe cases may have been overrepresented among those hospitalized, potentially leading to increased reimbursement. Conversely, it is also possible that the most severe cases died from COVID-19 before hospitalization for ACVD, resulting in a hospitalized population composed of less severe cases and, consequently, lower expenditures. In any case, a gradual return to pre-pandemic levels by 2022 and 2023 was anticipated, supported by the widespread vaccination campaign in 2021 and the progressive decline in the pandemic’s intensity over time, as reflected in the WHO’s decision to declare the end of the PHEIC in May 2023[20].

The objective of this study was to asses changes in healthcare utilization among patients hospitalized for ACVD during the pandemic period (2020–2023), as measured by global healthcare reimbursements.

## Methods

### Study Population

This open cohort study of the French population, conducted between January 1, 2015, and December 31, 2023, was reported following the STROBE guidelines [21]. Healthcare expenditures (in euros) were assessed from the perspective of the national health insurance system and included only reimbursed direct medical costs attributable to patient care. Throughout the study, the term “sex” referred to the sex assigned at birth. In line with the SAGER guidelines[22], sex was consistently taken into account in both the study design and reporting. Patients with at least one episode of ACVD, including heart failure, stroke, myocardial infarction, or pulmonary embolism, were included in the study cohort. Between 2020 and 2023, the ACVD population was further stratified based on whether patients had also been hospitalized for COVID-19 during the same year. A comparison with a control group was conducted to assess whether the observed changes in healthcare reimbursements were specific to ACVD.

### Data sources

All data were obtained from the French National Health Data System (Système National des Données de Santé, SNDS), which encompassed healthcare reimbursement information for nearly the entire French population, including data from the hospital discharge database [23]. Patients hospitalized for COVID-19 were defined as those who spent at least one night in the hospital with an ICD-10 code of U07.1, U07.10, U07.11, U07.14, or U07.15 recorded as the main or a related diagnosis [24]. Patients with ACVD were defined by using algorithms from the Healthcare Expenditures and Conditions Mapping (HECM)[25], based on hospital diagnosis codes according to the International Classification of Diseases, 10th Revision (ICD-10) (see Supplementary Text 1). Additionally, the HECM provided total annual healthcare expenditures per individual, categorized into 21 expenditure categories (see Supplementary Text 2). With the increasing interest in real-world data derived from large administrative healthcare databases, the HECM has already been used to assess the economic burden of diseases in France from 2015 to 2019[25]. The absence of data for a diagnosis or an event was considered to be the absence of an actual diagnosis or event, so that there was no missing data for analysis.

### Outcomes of interest

Global annual reimbursements for all healthcare consumption were analyzed among patients with at least one episode of ACVD during the year, including both ACVD-related and non–ACVD-related expenditures, along with their distribution across 21 expenditure categories.

### Comparative analysis

To assess whether the changes in healthcare expenditure reimbursement were specific to patients hospitalized for ACVD, a comparative analysis was performed using a control group not hospitalized for ACVD. This group of patients was matched on age, sex, COVID-19 status, and comorbidity severity in order to distinguish changes specifically associated with ACVD hospitalizations from those observed in the general population (see Supplementary Text 3).

### Statistical Analysis of Healthcare Expenditures

To quantify the impact of the pandemic period on healthcare expenditures across 21 categories, a two-step analytical approach was developed to compare expected and observed reimbursed amounts.

First, the pre-pandemic period (2015–2019) was used as a reference to model the evolution of healthcare expenditures prior to the onset of the COVID-19 pandemic. A linear regression model was applied to the average reimbursed amount per patient per year, which was obtained by dividing the total reimbursed amount in each stratum by the number of patients. Expected values for the years 2020 to 2023 were estimated for each expenditure category using the fitted models, under the assumption that pre-pandemic trends would have continued in the absence of the pandemic.

Second, observed and expected values for the pandemic years were compared to estimate deviations observed in the pandemic period. Similar modeling approaches had previously been used, for instance, to estimate excess mortality during the pandemic[26–28].

The regression model was adjusted for age class (in 5-year intervals, with 95–99+ for patients aged 95 and older), sex, calendar year (as a numeric variable), and the Mortality-Related Morbidity Index (MRMI)[29]. To account for potential differences in temporal dynamics across population strata, interaction terms between these variables were included. Further details on variable selection and interaction terms are provided in Supplementary Text 3 and Supplementary Figures 6 to 10. The regression used for modeling healthcare expenditures in each stratum was:

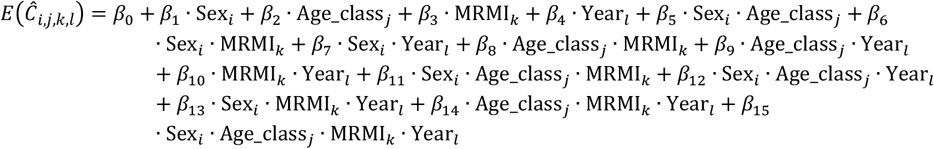

Let 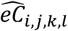denote the average reimbursed amount expected to occur according to the model in year *l* (*lϵ*{2020, 2021, 2022, 2023}) for the stratum defined by sex *i*, age class *j*, and comorbidity level *k*. The total expected reimbursed amount in each stratum, 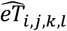,was obtained by multiplying 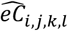 by the corresponding number of patients in that stratum. The observed reimbursed amount in the SNDS is denoted as 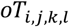.

The difference between observed and expected values was then computed as:

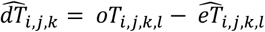

The deviation observed in year *l* relative to the expected evolution of the reimbursed amount was then calculated as follows, with all amounts expressed per patient-year:

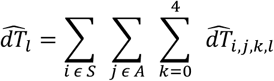

With *S* denoting the sex categories (males, females), *A* the 5-year age classes (0-4 to 95-99+ years), and K the comorbidity level (0 to 4+ MRMI).

All analyses were performed with statistical software R 4.0.2 (R Foundation for Statistical Computing, Vienna, Austria), the package stats was used for the general linear model, and 95% confidence intervals (CIs) were estimated with 1,000 bootstrap replications.

## Results

### Study population

Between 2015 and 2023, 3,899,712 ACVD-related patient-years were observed, with associated total expenditures of €85,791,476,762, corresponding to an average annual expenditure of €21,999 per patient. Females represented 45.7% of these patients and had an average expenditure of €21,119.

The median age of patients was 78 years [IQR: 66; 87], with a median of 83 years [72; 89] for females and 74 years [62; 84] for males.

### Evolution of healthcare expenditures

As shown in Figure 1, the average annual amount reimbursed per patient steadily increased during the pre-pandemic period, with yearly increments ranging from €94 to €215. However, a major increase of €1,007 was observed in 2021, followed by another substantial rise of €492 in 2022, and an even larger increase of €1,320 in 2023. Considering the 1,725,714 ACVD-related patient-years in years 2020 to 2023, 65,195 (3.8%) concerned patient-years with at least one COVID-19-related hospitalization, with an average annual expenditure of €38,300. Detailed annual information is presented in the flow diagram in Figure 2. A comprehensive interactive tool is provided in the Supplementary Digital Material to explore the full spectrum of health care reimbursements in France from 2015 to 2023 in the study population. This tool presents detailed annual data, including total reimbursed amounts, number of patients, gender distribution, comorbidity scores, and average reimbursement per patient. For the pandemic years, it also distinguishes between patients hospitalized for COVID-19 and those who were not. Reimbursements are further broken down into three major expenditure categories and 21 detailed categories. Users can dynamically filter the data by year, population subgroup, or type of expenditure to explore the evolution of health care reimbursements over time (see Supplementary Digital Material).

**Figure 1.**
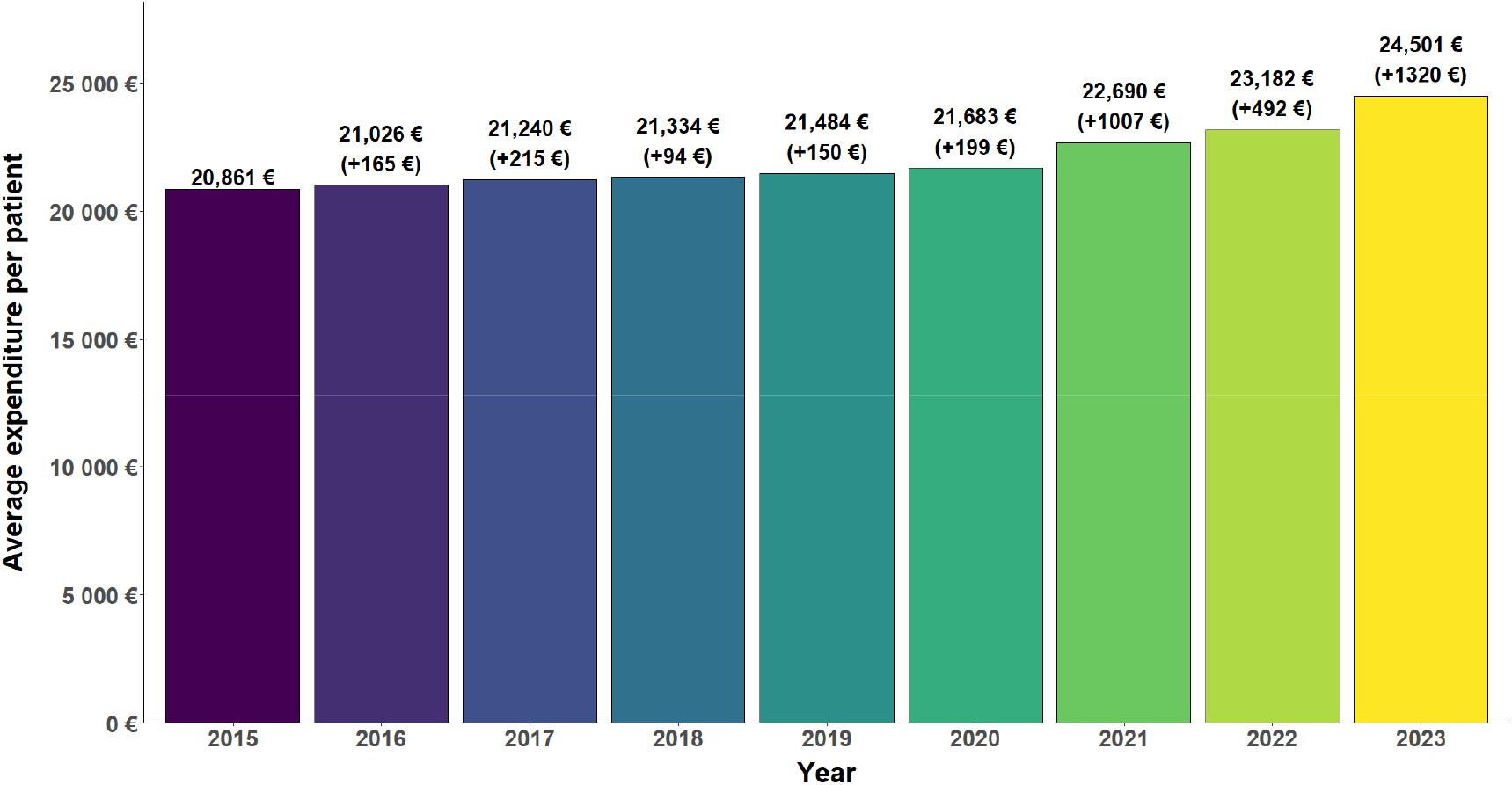
Average annual expenditure per ACVD patient, with rise compared to the previous year, France, 2015–2023. Each bar represents the annual average amount reimbursed per patient between 2015 and 2023. The number in brackets represents the difference in amount between year N and year N-1. For instance, in 2023 (yellow bar) the average annual amount reimbursed per patient hospitalized for ACVD was €24,501; representing an augmentation of €1320 compared to the average amount of €23,182 reimbursed in 2022.

**Figure 2.**
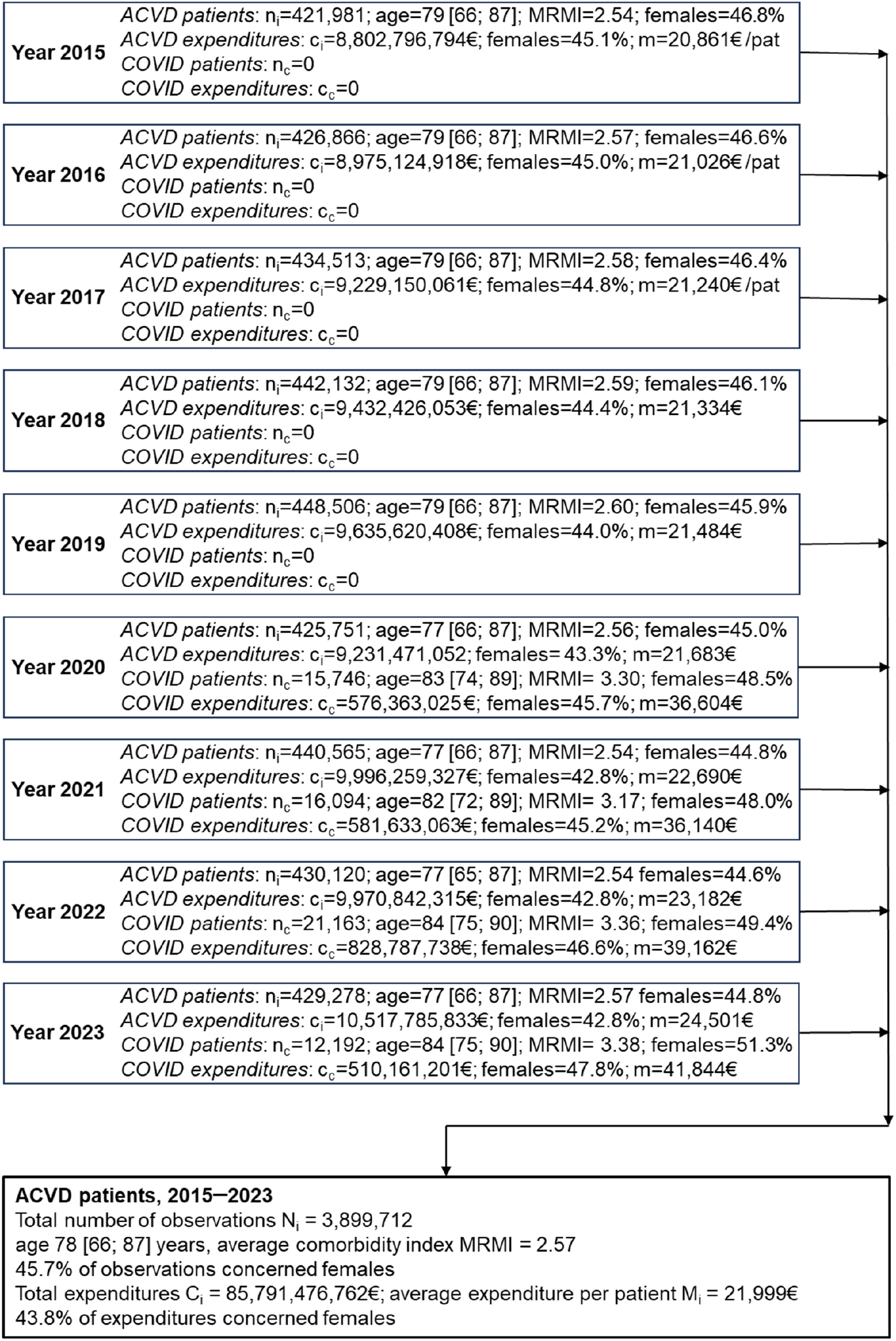
Flow diagram, France, 2015–2023. ACVD patient: Patient hospitalized for at least one ACVD within the year. COVID patient: patient with ACVD experiencing at least one COVID-19-related hospitalization. MRMI: Mortality-Related Morbidity Index. In each year-specific cell, n corresponds to the number of patients observed this given year (also equivalent to the number of patient-years observed this given year) and c to the corresponding annual expenditures, including all healthcare expenditures. Age is reported as the median value [interquartile range], and the percentage of data concerning females is also reported.

As detailed in Table 1, the expenditures observed from 2020 to 2023, following the onset of the pandemic, consistently remained above the expected expenditures based on the trend from the five pre-pandemic years (2015–2019) for both males and females. The excess worsened over time, starting with €51,266,040 (95% CI: €48,855,492; €53,506,955) more than expected in 2020 and peaking at €1,109,888,907(€1,106,424,348; €1,113,980,482) in 2023. These estimates correspond to annual increases ranging from 0.6% to 11.9%. Over the entire pandemic period, there were €2,298,166,970 (€2,292,788,304; €2,303,326,450) excess expenditure, representing a 6.2% increase from the expected figures. Males were more concerned each year, with a total excess expenditure of €1,422,478,782 (€1,418,869,707; €1,426,874,217) compared to €875,688,187 (€871,663,811; €881,259,859) for females. Patients with an MRMI of 3 or more accumulated an excess expenditure of €1,368,873,629 (€1,363,771,120; €1,373,307,964) between 2020 and 2023, while they represented 43.6% of the patients.

**Table 1:**
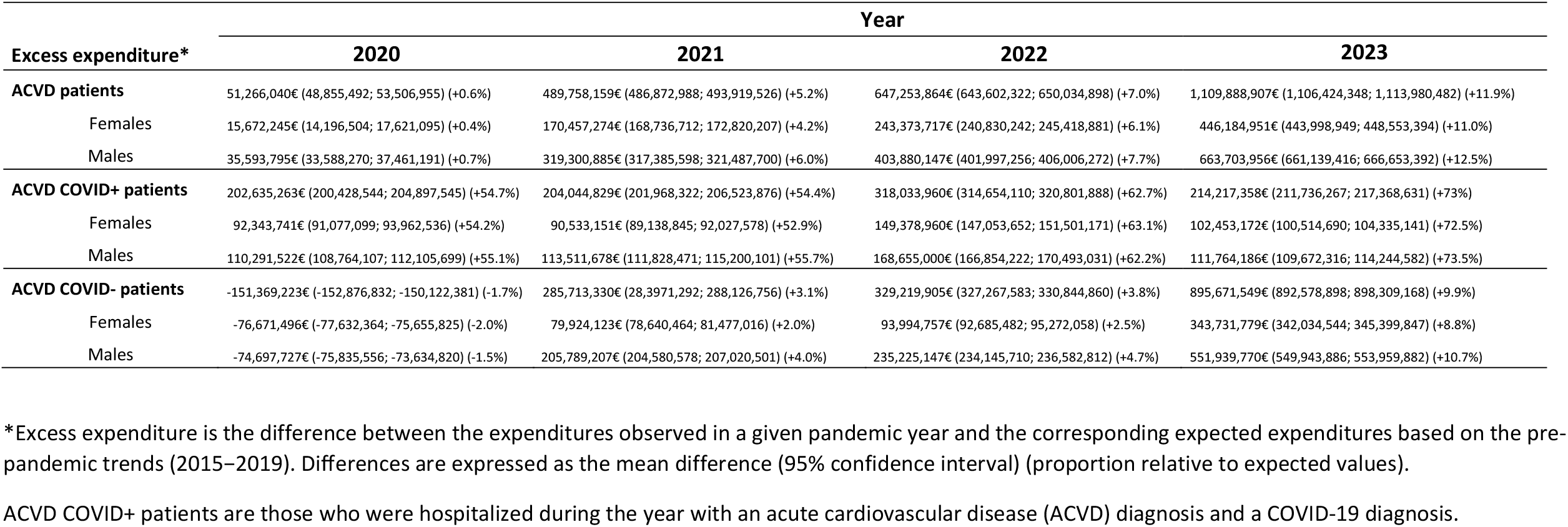
Estimated excess expenditure, ACVD patients and ACVD + COVID patients, France, 2020*−*2023.

### Evolution of healthcare expenditures of patients with ACVD hospitalized for COVID-19

Although the 65,195 patient-years observed for patients with ACVD who also experienced COVID-19-related hospitalization(s) between 2020 and 2023 represented only 3.8% of the effectives, they accounted for a substantial excess expenditure of €938,931,410 (€934,539,400 to €944,574,066).

This corresponds to a 60.7% increase (60.5% to 60.9%) compared to the expected value and represents 40.9% of the total excess expenditure during the pandemic years. This amount rose from €202,635,263 (€200,428,544; €204,897,545) in 2020 to €318,033,960 (€314,654,110; €320,801,888) in 2022 before decreasing to €214,217,358 (€211,736,267; €217,368,631) in 2023. These estimates correspond to annual increases ranging from 54.4% to 73.0%. These increases affected males and females equally (see Table 1).

### Excess expenditure by healthcare expense category

As illustrated in Figure 3, excess healthcare expenditure for patients with ACVD who also experienced COVID-19-related hospitalization(s) primarily concerned two healthcare expense categories: short-stay hospitalizations and hospitalizations in rehabilitation care accounted for 89.6% of the corresponding global economic burden, with an estimated excess expense of €841,505,828 (€836,741,679; €846,865,022) between 2020 and 2023. Though this accounted for a limited amount, expenditures for hospitalizations in psychiatry substantially increased during pandemic years, from 34% to 86% as compared to expected values (see Supplementary Figures 11 and 12).

**Figure 3.**
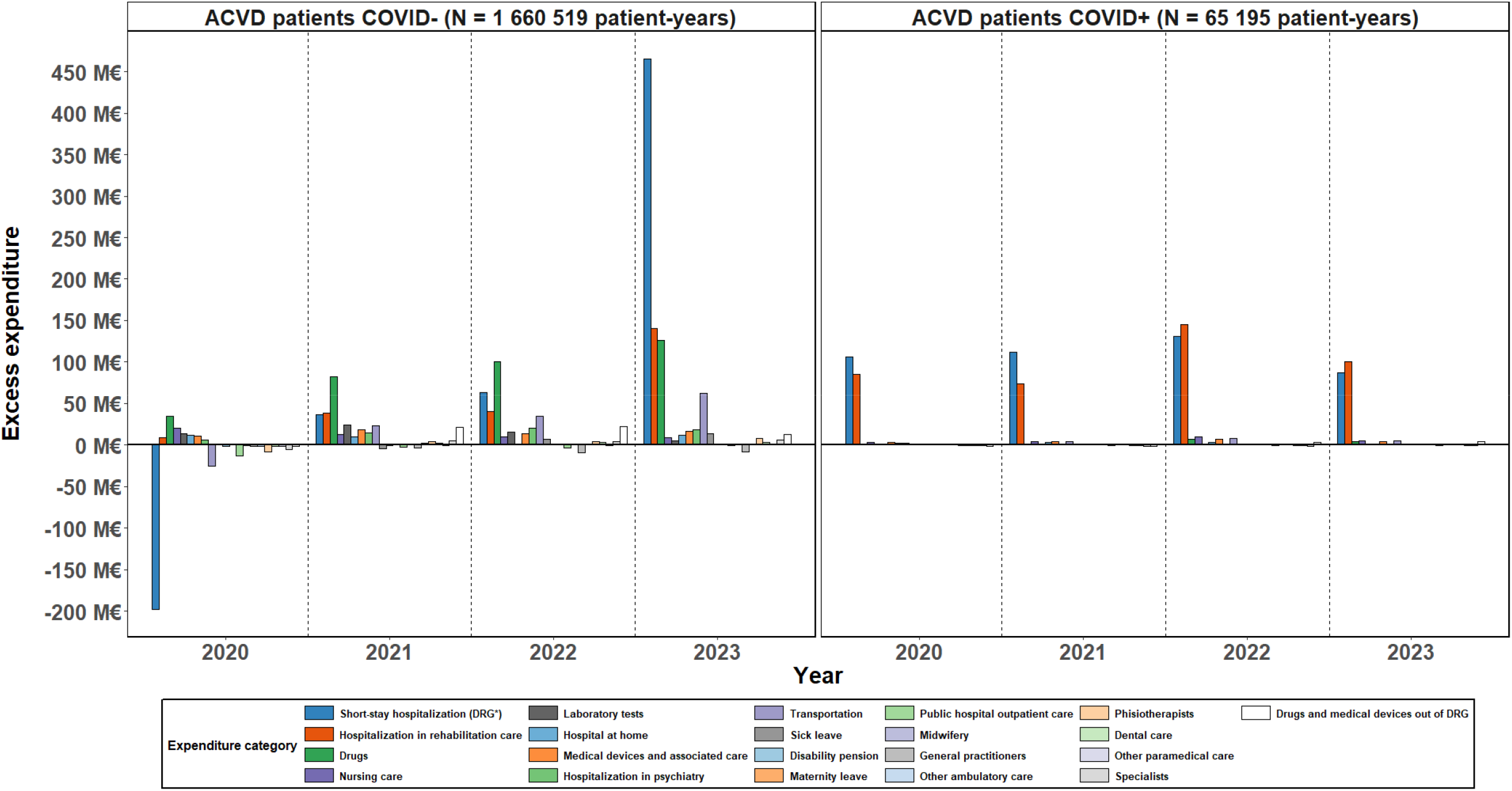
Observed minus expected expenditure per category and COVID-19 status, ACVD patients, France, 2020–2023. *DRG: diagnosis-related group Annual difference between observed and expected reimbursed amounts, by year, expenditure category, and COVID-19 status, for patients hospitalized for ACVD. For example, in 2020, among patients hospitalized for ACVD and for COVID-19 (right panel), expenditures related to short-stay hospitalizations (dark blue bar) exceeded expected values (based on 2015–2019 trends) by more than €100 million.

For patients not hospitalized for COVID-19, several disturbances occurred during the pandemic period (see Figure 3). Laboratory tests and drugs expenses exceeded expected values for three consecutive years, with excesses ranging from 5% to 33%. Hospitalizations in rehabilitation care also exceeded expectations, as did hospitalizations in psychiatry. Regarding short-stay hospitalizations, a 4% decrease in reimbursements was observed in 2020, followed by a slight increase of 1% in both 2021 and 2022, and a sharp increase of 10% in 2023 (see Supplementary Figures 13 and 14).

## Discussion

### Key results

In this cohort study of patients with ACVD in France from 2015 to 2023, accounting for 3.9 million patient-years, the evolving trends in healthcare expenditures were investigated in detail to quantify the extent of disruption during the COVID-19 pandemic period (2020–2023). Analyses by age, sex, COVID-19 status, year, comorbidity score, and expenditure categories were performed to identify the patients and categories most affected. A major and prolonged disruption has been highlighted since the onset of the pandemic, persisting through 2023. Specifically, healthcare expenditures rose by 0.6% in 2020 and increased to 11.9% in 2023, resulting in a global economic burden of €2.3 billion. Importantly, 41% of this burden was driven by ACVD patients with COVID-19-related hospitalization(s), although they represented only 4% of the corresponding patient-years taken into account for calculating the excess expenditure during the pandemic years. Males and patients with high comorbidity levels particularly contributed to this burden. Overall, ACVD patients experiencing COVID-19-related hospitalization(s) had an average annual excess expenditure of €14,402, while other ACVD patients had an excess expenditure of €1,384. Regarding expenditure categories, the majority of the excess cost was driven by short-stay hospitalizations and hospitalizations in rehabilitation care among ACVD patients with COVID-19-related hospitalization(s), and a marked relative increase was also observed in hospitalizations in psychiatry. Among other ACVD patients who were not hospitalized for COVID-19, significant disruptions in healthcare reimbursements were observed across short-stay hospitalizations, rehabilitation care, and drug consumption.

Similar to the ACVD population, excess expenditures were also observed in the control group throughout the entire pandemic period, with the majority driven by patients who experienced COVID-19-related hospitalization(s), and involving similar expenditure categories. However, between 2020 and 2023, the total increase amounted to 10.9% in the control group, compared to 6.2% in the ACVD population (see Supplementary Figures 1–5 and Supplementary Table 1).

### Interpretation

Expenditures related to the prevention or management of COVID-19 infections could account for a major part of the economic burden. Indeed, patients with ACVD who also experienced COVID-19 hospitalization(s) represented a relatively small group, yet they accounted for a substantial portion of the overall burden. Among these patients, short-stay hospitalizations, probably to handle the acute phase of the disease, recorded the highest global increase. Indeed, COVID-19 patients often require intensive care with respiratory devices, leading to high related costs[30, 31]. A systematic review on the economic burden of COVID-19 highlighted that the main drivers for higher costs were ICU admission and in-hospital resource use, such as mechanical ventilation[4]. After the acute phase, COVID-19 patients required rehabilitation care, which aligns with the substantially increased cost in rehabilitation hospitalizations [32, 33]. Furthermore, our study highlighted that males and patients with a high number of comorbidities highly contributed to excess expenditure, a profile consistent with known COVID-19 risk factors[34].

Among ACVD patients not hospitalized for COVID-19, the effects of the pandemic period might also explain rises in some expenditure categories. For instance, some increase in expenses for medication is likely due to the widespread adoption of preventive measures such as COVID-19 self-tests and protective masks. Laboratory expenditures were also impacted by the extensive screening policies implemented during the pandemic. Notably, in France, €1.9 billion, €300 million, and €100 million were respectively spent on laboratory tests, antigenic tests, and protective masks in 2020 [35]. However, the direct effects of the pandemic period cannot fully explain all observed disruptions. For example, increases in expenditures for hospitalizations in psychiatry among patients with ACVD concerned both those who had and those who had not experienced COVID-19-related hospitalization(s), strongly suggesting that the corresponding excess expenditures were more likely indirect consequences. These may include the anxiety-inducing context of a new pandemic, major changes such as lockdowns and containment measures, uncertainty about employment, and fear of contamination. These indirect consequences have led to detrimental mental health outcomes[36, 37] and may have increased hospitalizations in psychiatry and drug consumption[38–41]. With the widespread vaccination campaigns in 2021[42, 43] and the decreasing intensity of the pandemic, marked by lower COVID-19 mortality over time[44], health expenditures were expected to progressively return to pre-pandemic trends by 2023, particularly for patients not hospitalized for COVID-19. However, our study indicates that excess expenditure in patients with ACVD continued to rise in 2023. This persistent disruption suggests that the pandemic period has had critical and lasting impacts on the management of patients with ACVD.

The study focuses on patients with at least one hospitalization for ACVD and accounts for all annual healthcare expenditures, regardless of their direct association with the ACVD condition. However, publicly available data from the French National Health Insurance indicate a noticeable increase in average healthcare expenditures related to these diseases between 2020 and 2023[45–48]. This rise, which marks a clear rupture from pre-pandemic trends, aligns with our findings and supports the validity of the expenditure patterns observed in this study.

One contributing factor to the excess cost observed in short-stay hospitalizations may be the increase in hospitalization tariffs, which was particularly pronounced in 2023[49]. Tariff levels rose sharply during the pandemic period[50], in contrast to previous years, partly due to the “Ségur de la santé”[51] reform implemented by the French government in response to the COVID-19 crisis.

Between 2020 and 2023, the cumulative increase in hospitalization tariffs exceeded 10%, which is consistent with the rise observed in 2023 for this sector.

Although patients with ACVD experienced significant excess expenditures throughout the entire pandemic period, the increase was nearly twice as high in the control group. This discrepancy may reflect the positive impact of protective measures or specific care adaptations implemented for patients with ACVD during the pandemic.

### Generalizability of the results

Study results stand for France and cannot be directly extended to other countries. Nevertheless, disruptions in healthcare utilizations and expenditure have been reported in other countries[52–55], suggesting that the broader implications of the study may still be relevant.

### Strengths and limitations

One of the primary strengths of this study is the exhaustiveness of the data covering almost the entire French population over nine years. Based on a medico-administrative database originally designed for healthcare reimbursement purposes, the comprehensive dataset used in this study ensures its representativeness of the population, enabling a reliable and precise estimation of the pandemic period’s impact on healthcare consumption and associated expenditures. The extensive temporal scope allows for a robust analysis of trends and variations in healthcare utilization and costs, documenting in detail the pandemic’s effects over time.

Nevertheless, this study also has several limitations. As with any study based on medico-administrative databases, coding errors cannot be excluded, particularly during crisis periods such as the COVID-19 pandemic. These errors may affect the identification of relevant pathologies and the definition of various expense categories. However, given the large sample sizes and the magnitude of the estimated disruptions, potential coding errors are unlikely to alter the overall conclusions of the study. Another limitation pertains to the measurement of economic impact. While the study considers the full range of healthcare expenditures across 21 categories, indirect economic impacts, such as productivity losses due to work absences[56], were not investigated.

## Conclusion

While acute cardiovascular diseases require rapid management, and corresponding care pathways were theoretically maintained during the pandemic, our findings highlighted a major and persistent disruption in healthcare reimbursements starting in early 2020 and further intensifying in 2023. A substantial portion of these excess costs appears to be directly linked to severe SARS-CoV-2 infections and pandemic-related prevention measures. However, increasing disruptions were also observed among patients not hospitalized for COVID-19 and in non-specific areas such as psychiatry. Strikingly, even greater disruptions were estimated in the control group, suggesting that the broader healthcare system has been impacted. These findings are of critical importance for public health, not only to prepare for future crises but also to better understand and mitigate ongoing healthcare system disruptions in the management of this vulnerable population.

## Supporting information

Supplementary Digital Material

## Ethics approval and consent to participate

The SNDS is a set of strictly anonymous databases comprising all mandatory national health insurance reimbursement data. Since June 30, 2021, INSERM has direct access to the SNDS. This permanent access is provided according French Decree No. 2016-1871 of December 26, 2016 relating to the processing of personal SNDS data and French law articles Art. R. 1461-1325 and 14. This study was declared prior to its initiation to the CepiDc-INSERM registry of studies requiring the use of the SNDS. In accordance with national legislation and EU General Data Protection Regulation, written informed consent for participation was not required for this study.

## Availability of data and materials

According to the principles of data protection and French regulations, the authors cannot publicly release the data from the French National Health Data System (SNDS). However, any person or structure, public or private, for-profit or non-profit, can access SNDS data upon authorisation from the French Data Protection Office (CNIL, Commission Nationale de l’Informatique et des Libertés) to carry out a study, research, or an evaluation of public interest (https://www.snds.gouv.fr/SNDS/Contexte-et-perspectives-reglementaires#).

## Competing Interests

The authors of this manuscript have no conflict of interest to disclose.

## Funding

This work was supported by the Initiative Économie de la Santé of Sorbonne Université (Idex Sorbonne Université, programmes Investissements d’Avenir), and by the Ministère de la Solidarité et de la Santé (Programme de Recherche sur la Performance du Système des Soins, PREPS 20-0163). The sponsor and the funders had no role in study design, data collection and analysis, interpretation of data, decision to publish, or preparation of the manuscript.

## Author Contributions

TD, GH, and NL initiated the study. GH and NL supervised the study; GH, NL, and PM designed the experimental plan; PM managed data and performed the analyses; GH, NL, and PM can take responsibility for the integrity of the data and the accuracy of the data analysis, PM is the guarantor; GH, NL, and PM prepared the first draft of the manuscript; All authors (TD, ME, GH, MK, SLC, NL, PM, and AR) contributed to interpretation of the data, critically revised the manuscript, and approved the final version.

## Acknowledgements

The COVID-HOSP study group: Tristan Delory (Centre Hospitalier Annecy Genevois, Annecy, France); Fanny Duchaine (IRDES, Paris, France); Maude Espagnacq (IRDES, Paris, France); Gilles Hejblum (INSERM, Paris, France); Myriam Khlat (INED, Aubervilliers, France); Nathanaël Lapidus (INSERM, Paris, France); Sophie Le Cœur (INED, Aubervilliers, France); Elhadji Leye (INSERM, Paris, France); Paul Moulaire (INSERM, Paris FRANCE); Jonas Poucineau (INED, Aubervilliers, France).

## Notes

### Competing Interest Statement

The authors have declared no competing interest.

### Funding Statement

This work was supported by the Initiative Economie de la Sante of Sorbonne Universite (Idex Sorbonne Universite, programmes Investissements d'Avenir), and by the Ministere de la Solidarite et de la Sante (Programme de Recherche sur la Performance du Systeme des Soins, PREPS 20-0163). The sponsor and the funders had no role in study design, data collection and analysis, interpretation of data, decision to publish, or preparation of the manuscript.

### Author Declarations

The SNDS is a set of strictly anonymous databases comprising all mandatory national health insurance reimbursement data. Since June 30, 2021, INSERM has direct access to the SNDS. This permanent access is provided according French Decree No. 2016-1871 of December 26, 2016 relating to the processing of personal SNDS data and French law articles Art. R. 1461-1325 and 14. This study was declared prior to its initiation to the CepiDc-INSERM registry of studies requiring the use of the SNDS.

## References

1. Maxmen A. Wuhan market was epicentre of pandemic’s start, studies suggest. Nature. 2022;603:15–6. 10.1038/d41586-022-00584-8.

2. World Health Organization. Statement on the second meeting of the International Health Regulations (2005) Emergency Committee regarding the outbreak of novel coronavirus (2019-nCoV). https://www.who.int/news/item/30-01-2020-statement-on-the-second-meeting-of-the-international-health-regulations-(2005)-emergency-committee-regarding-the-outbreak-of-novel-coronavirus-(2019-ncov). Accessed 11 Sep 2024.

3. Wang H, Paulson KR, Pease SA, Watson S, Comfort H, Zheng P, et al. Estimating excess mortality due to the COVID-19 pandemic: a systematic analysis of COVID-19-related mortality, 2020–21. The Lancet. 2022;399:1513–36. 10.1016/S0140-6736(21)02796-3.

4. Richards F, Kodjamanova P, Chen X, Li N, Atanasov P, Bennetts L, et al. Economic Burden of COVID-19: A Systematic Review. Clin OUTCOMES Res. 2022;14:293–307. 10.2147/CEOR.S338225.

5. Leal J, Luengo-Fernández R, Gray A, Petersen S, Rayner M. Economic burden of cardiovascular diseases in the enlarged European Union. Eur Heart J. 2006;27:1610–9. 10.1093/eurheartj/ehi733.

6. Luengo-Fernandez R, Walli-Attaei M, Gray A, Torbica A, Maggioni AP, Huculeci R, et al. Economic burden of cardiovascular diseases in the European Union: a population-based cost study. Eur Heart J. 2023;44:4752–67. 10.1093/eurheartj/ehad583.

7. Nicolas M, Pestel L, Rivière S, Rachas A, Gastaldi-Menager C. Economic burden of cardiovascular diseases from 2012 to 2017 based on French national claim database. Eur J Public Health. 2019;29 Supplement_4:ckz187.069. 10.1093/eurpub/ckz187.069.

8. Khan Y, Verhaeghe N, Devleesschauwer B, Cavillot L, Gadeyne S, Pauwels N, et al. The impact of the COVID-19 pandemic on delayed care of cardiovascular diseases in Europe: a systematic review. Eur Heart J Qual Care Clin Outcomes. 2023;9:647–61. 10.1093/ehjqcco/qcad051.

9. Kiss P, Carcel C, Hockham C, Peters SAE. The impact of the COVID-19 pandemic on the care and management of patients with acute cardiovascular disease: a systematic review. Eur Heart J - Qual Care Clin Outcomes. 2021;7:18–27. 10.1093/ehjqcco/qcaa084.

10. Olié V, Isnard R, Pousset F, Grave C, Blacher J, Gabet A. Epidemiology of hospitalized heart failure in France based on national data over 10 years, 2012–2022. ESC Heart Fail. n/a n/a. 10.1002/ehf2.15137.

11. Gabet A, Grave C, Tuppin P, Chatignoux E, Béjot Y, Olié V. Impact of the COVID-19 pandemic and a national lockdown on hospitalizations for stroke and related 30-day mortality in France: A nationwide observational study. Eur J Neurol. 2021;28:3279–88. 10.1111/ene.14831.

12. Grave C, Gabet A, Puymirat E, Empana J-P, Tuppin P, Danchin N, et al. Myocardial infarction throughout 1 year of the COVID-19 pandemic: French nationwide study of hospitalization rates, prognosis and 90-day mortality rates. Arch Cardiovasc Dis. 2021;114:768–80. 10.1016/j.acvd.2021.10.008.

13. Gabet A, Grave C, Tuppin P, Emmerich J, Olié V. Changes in the epidemiology of patients hospitalized in France with deep venous thrombosis and pulmonary embolism during the COVID-19 pandemic. Thromb Res. 2021;207:67–74. 10.1016/j.thromres.2021.09.009.

14. Chouairi F, Pinsker B, Miller PE, Fudim M. Effects of COVID-19 on heart failure admissions. Am Heart J. 2023;263:183–7. 10.1016/j.ahj.2023.05.001.

15. Chan K, Baker J, Conroy A, Rubens M, Ramamoorthy V, Saxena A, et al. Burden of cardiovascular disease on coronavirus disease 2019 hospitalizations in the USA. Coron Artery Dis. 2024;35:584–9. 10.1097/MCA.0000000000001390.

16. Aghdam BH, Seyedbaglou ZK, Akhtari AS. Comparison of Mortality, Length of Stay, and Hospitalization Costs of Hospitalized COVID-19 Patients with Cardiac and Non-Cardiac Disease. Open J Emerg Med. 2023;11:57–67. 10.4236/ojem.2023.113007.

17. Leboucher C, Blein C, Machuron V, Benyounes K, Le lay K, Millier A, et al. The burden of hospitalisations for COVID-19 in France: a study of all cases in the national insurance claims database in 2020. J Mark Access Health Policy. 2023;11:2160328. 10.1080/20016689.2022.2160328.

18. Demont C, Mouaddin NE, Chillotti L, Uhart M, Chéret A. Economic Burden of Influenza and COVID-19 in French Hospitals: Analysis of PMSI Data From 2018 to 2022. Value Health. 2024;27:S588–9. 10.1016/j.jval.2024.10.3640.

19. Yang J, Tamberou Cheikh, Arnee, Elise, Squara, Pierre-Alexandre, Boukhlal, Ayoub, Nguyen, Jennifer L., et al. All-cause healthcare resource utilization and costs among community-managed adults with long-COVID in France, 2020–2023. J Med Econ. 2025;28:535–43. 10.1080/13696998.2025.2485626.

20. World Health Organization. Statement on the fifteenth meeting of the IHR (2005) Emergency Committee on the COVID-19 pandemic. https://www.who.int/news/item/05-05-2023-statement-on-the-fifteenth-meeting-of-the-international-health-regulations-(2005)-emergency-committee-regarding-the-coronavirus-disease-(covid-19)-pandemic. Accessed 28 Jun 2024.

21. Vandenbroucke JP, Elm E von, Altman DG, Gøtzsche PC, Mulrow CD, Pocock SJ, et al. Strengthening the Reporting of Observational Studies in Epidemiology (STROBE): Explanation and Elaboration. PLOS Med. 2007;4:e297. 10.1371/journal.pmed.0040297.

22. Heidari S, Babor TF, De Castro P, Tort S, Curno M. Sex and Gender Equity in Research: rationale for the SAGER guidelines and recommended use. Res Integr Peer Rev. 2016;1:2. 10.1186/s41073-016-0007-6.

23. Tuppin P, Rudant J, Constantinou P, Gastaldi-Menager C, Rachas A, de Roquefeuil L, et al. Value of a national administrative database to guide public decisions: From the systeme national d’information interregimes de l’Assurance Maladie (SNIIRAM) to the systeme national des donnees de sante (SNDS) in France. Rev Epidemiol Sante Publique. 2017;65:S149–67. 10.1016/j.respe.2017.05.004.

24. Méthode. https://www.assurance-maladie.ameli.fr/etudes-et-donnees/par-theme/pathologies/cartographie-assurance-maladie/methode-cartographie-pathologies-depenses-assurance-maladie. Accessed 18 Feb 2025.

25. Rachas A, Gastaldi-Ménager C, Denis P, Barthélémy P, Constantinou P, Drouin J, et al. The Economic Burden of Disease in France From the National Health Insurance Perspective. Med Care. 2022;60:655–64. 10.1097/MLR.0000000000001745.

26. Alicandro G, La Vecchia C, Islam N, Pizzato M. A comprehensive analysis of all-cause and cause-specific excess deaths in 30 countries during 2020. Eur J Epidemiol. 2023;38:1153–64. 10.1007/s10654-023-01044-x.

27. Barnard S, Chiavenna C, Fox S, Charlett A, Waller Z, Andrews N, et al. Methods for modelling excess mortality across England during the COVID-19 pandemic. Stat Methods Med Res. 2022;31:1790–802. 10.1177/09622802211046384.

28. Moulaire P, Hejblum G, Lapidus N. Excess mortality and years of life lost from 2020 to 2023 in France: a cohort study of the overall impact of the COVID-19 pandemic on mortality. BMJ Public Health. 2025;3. 10.1136/bmjph-2024-001836.

29. Constantinou P, Tuppin P, Fagot-Campagna A, Gastaldi-Ménager C, Schellevis FG, Pelletier-Fleury N. Two morbidity indices developed in a nationwide population permitted performant outcome-specific severity adjustment. J Clin Epidemiol. 2018;103:60–70. 10.1016/j.jclinepi.2018.07.003.

30. Yehoshua A, Cook, Angela D., Di Fusco, Manuela, Rudolph, Abby E., Thoburn, Elizabeth, Lopez, Santiago M.C, et al. Health outcomes and economic burden among patients with a COVID-19-associated hospitalization in the United States during the predominance of the XBB and JN.1 omicron lineages. J Med Econ. 2024;27:1372–8. 10.1080/13696998.2024.2416873.

31. Di Fusco M, Shea Kimberly M., Lin Jay, Nguyen, Jennifer L., Angulo Frederick J., Benigno, Michael, et al. Health outcomes and economic burden of hospitalized COVID-19 patients in the United States. J Med Econ. 2021;24:308–17. 10.1080/13696998.2021.1886109.

32. Gallien S, Guilmet C, Hurtes A, Omichessan H, Messaoudi F, Abou chakra C, et al. Coûts des hospitalisations et des soins de suite et de réadaptation liés au COVID-19 en France en 2020. Médecine Mal Infect Form. 2022;1:S51. 10.1016/j.mmifmc.2022.03.110.

33. Ong PL, Tay MRJ, Tham SL. Cost of Rehabilitation in Critically Ill COVID-19 Survivors: A Little Goes a Long Way. J Int Soc Phys Rehabil Med. 2021;4:104. 10.4103/JISPRM-000129.

34. Semenzato L, Botton J, Drouin J, Cuenot F, Dray-Spira R, Weill A, et al. Chronic diseases, health conditions and risk of COVID-19-related hospitalization and in-hospital mortality during the first wave of the epidemic in France: a cohort study of 66 million people. Lancet Reg Health - Eur. 2021;8:100158. 10.1016/j.lanepe.2021.100158.

35. DREES. Les dépenses de prévention 2022.

36. Global prevalence and burden of depressive and anxiety disorders in 204 countries and territories in 2020 due to the COVID-19 pandemic. Lancet Lond Engl. 2021;398:1700–12. 10.1016/S0140-6736(21)02143-7.

37. Xiong J, Lipsitz O, Nasri F, Lui LMW, Gill H, Phan L, et al. Impact of COVID-19 pandemic on mental health in the general population: A systematic review. J Affect Disord. 2020;277:55–64. 10.1016/j.jad.2020.08.001.

38. Fond G, Pauly V, Brousse Y, Llorca P-M, Cortese S, Rahmati M, et al. Mental Health Care Utilization and Prescription Rates Among Children, Adolescents, and Young Adults in France. JAMA Netw Open. 2025;8:e2452789. 10.1001/jamanetworkopen.2024.52789.

39. Lamer A, Saint-Dizier C, Ayed E, Levaillant M, Hamel J-F, Chazard E, et al. Augmentation persistante de la consommation de psychotropes chez les jeunes femmes suite à la pandémie de COVID-19. J Epidemiol Popul Health. 2024;72:202282. 10.1016/j.jeph.2024.202282.

40. Paitraud D. COVID19: la consommation d’anxiolytiques et d’antidépresseurs en forte hausse. VIDAL. 2021. https://www.vidal.fr/actualites/27177-covid-19-la-consommation-d-anxiolytiques-et-d-antidepresseurs-en-forte-hausse.html. Accessed 28 Jul 2025.

41. Tiger M, Castelpietra G, Wesselhoeft R, Lundberg J, Reutfors J. Utilization of antidepressants, anxiolytics, and hypnotics during the COVID-19 pandemic. Transl Psychiatry. 2024;14:175. 10.1038/s41398-024-02894-z.

42. Meslé MM, Brown J, Mook P, Hagan J, Pastore R, Bundle N, et al. Estimated number of deaths directly averted in people 60 years and older as a result of COVID-19 vaccination in the WHO European Region, December 2020 to November 2021. Eurosurveillance. 2021;26:2101021. 10.2807/1560-7917.ES.2021.26.47.2101021.

43. SPF. Couverture vaccinale contre la Covid-19 et impact sur la dynamique de l’épidémie. https://www.santepubliquefrance.fr/import/couverture-vaccinale-contre-la-covid-19-et-impact-sur-la-dynamique-de-l-epidemie. Accessed 8 Apr 2025.

44. Fouillet A, Aubineau Y, Godet F, Costemalle V, Coudin E. Leading causes of death in France in 2023 and recent trends. https://beh.santepubliquefrance.fr/beh/2025/13/2025_13_1.html. Accessed 28 Jul 2025.

45. Caisse Nationale d’Assurance Maladie (CNAM). Pathologie — Data ameli—AVC. https://data.ameli.fr/pages/pathologies/?refine.patho_niv1=Maladies%20cardioneurovasculaires&refine.patho_niv2=Accident%20vasculaire%20c%C3%A9r%C3%A9bral&refine.patho_niv3=Accident%20vasculaire%20c%C3%A9r%C3%A9bral%20aigu. Accessed 28 May 2025.

46. Caisse Nationale d’Assurance Maladie (CNAM). Pathologie — Data ameli—EP. https://data.ameli.fr/pages/pathologies/?refine.patho_niv1=Maladies%20cardioneurovasculaires&refine.patho_niv2=Embolie%20pulmonaire. Accessed 28 May 2025.

47. Caisse Nationale d’Assurance Maladie (CNAM). Pathologie — Data ameli—ICA. https://data.ameli.fr/pages/pathologies/?refine.patho_niv1=Maladies%20cardioneurovasculaires&refine.patho_niv2=Insuffisance%20cardiaque&refine.patho_niv3=Insuffisance%20cardiaque%20aigu%C3%AB. Accessed 28 May 2025.

48. Caisse Nationale d’Assurance Maladie (CNAM). Pathologie — Data ameli—SCO. https://data.ameli.fr/pages/pathologies/?refine.patho_niv1=Maladies%20cardioneurovasculaires&refine.patho_niv2=Maladie%20coronaire&refine.patho_niv3=Syndrome%20coronaire%20aigu. Accessed 28 May 2025.

49. FHPMCO. Dépêche EXPERT N°740 - Arrêté Tarifaire 2023 / Reprise de la facturation. FHP-MCO. 2023. https://www.fhpmco.fr/2023/03/31/depeche-expert-n740-arrete-tarifaire-2023-reprise-de-la-facturation/. Accessed 28 Jul 2025.

50. Tarifs MCO et HAD | Publication ATIH. https://www.atih.sante.fr/tarifs-mco-et-had. Accessed 28 Jul 2025.

51. A D, A D. Ségur de la santé: les conclusions. Ministère du Travail, de la Santé, des Solidarités et des Familles. https://sante.gouv.fr/systeme-de-sante/segur-de-la-sante/article/segur-de-la-sante-les-conclusions. Accessed 28 Jul 2025.

52. Ramadani RV, Svensson M, Hassler S, Hidayat B, Ng N. Effects of the COVID-19 pandemic on healthcare utilization among older adults with cardiovascular diseases and multimorbidity in Indonesia: an interrupted time-series analysis. BMC Public Health. 2024;24:71. 10.1186/s12889-023-17568-6.

53. Haidar A, Gajjar A, Parikh RV, Benharash P, Fonarow GC, Watson K, et al. National Costs for Cardiovascular-Related Hospitalizations and Inpatient Procedures in the United States, 2016 to 2021. Am J Cardiol. 2025;234:63–70. 10.1016/j.amjcard.2024.10.003.

54. Dale CE, Takhar R, Carragher R, Katsoulis M, Torabi F, Duffield S, et al. The impact of the COVID-19 pandemic on cardiovascular disease prevention and management. Nat Med. 2023;29:219–25. 10.1038/s41591-022-02158-7.

55. Ascandar N, Chervu N, Bakhtiyar SS, Cho NY, Kim S, Orellana M, et al. Clinical and financial outcomes of hospitalizations for cardiac device infection during the COVID-19 pandemic in the US. PLOS ONE. 2023;18:e0291774. 10.1371/journal.pone.0291774.

56. Niewiadomski P, Ortega-Ortega M, Łyszczarz B. Productivity Losses due to Health Problems Arising from COVID-19 Pandemic: A Systematic Review of Population-Level Studies Worldwide. Appl Health Econ Health Policy. 2025;23:231–51. 10.1007/s40258-024-00935-8.

