## Supplementary Material for "Health care resource consumption and corresponding economic burden in patients with acute cardiovascular diseases during the COVID-19 pandemic period"

### Supplementary Text 1. Algorithms used to define Acute Cardiovascular Diseases (ACVD)

The algorithms of the Healthcare Expenditures and Conditions Mapping (HECM) G12 version[1] were used to define ACVD. These algorithms underwent external expert review[1] and had already been used in a previous study[2]. The corresponding ICD-10 codes and data sources are detailed below:

#### Acute Cardiovascular Diseases (MCV_CAT_AIG)

This group includes patients receiving care for Acute Coronary Syndrome (MCV_SCO_AIG), Acute Stroke (MCV_AVC_AIG), Acute Heart Failure (MCV_ICA_AIG), or Acute Pulmonary Embolism (MCV_EPU_IND).

#### Acute Coronary Syndrome (MCV_SCO_AIG)

This category includes patients hospitalized in short-stay medical and surgical units (MCO) during year n for acute ischemic heart disease, identified through the primary diagnosis (PD) of one or more hospital stays (RUMs).

ICD-10 codes used:

- I200+0 – Unstable angina with elevated myocardial biochemical markers (enzymes)
- I21 – Acute myocardial infarction
- I22 – Subsequent myocardial infarction
- I23 – Certain current complications following acute myocardial infarction
- I24 – Other acute ischemic heart diseases

#### Acute Stroke (MCV_AVC_AIG)

This group includes patients hospitalized during year n for acute cerebrovascular diseases, based on the primary diagnosis (PD) of one or more hospital stays (RUMs), excluding occlusions and stenoses of cerebral and precerebral arteries not resulting in cerebral infarction.

In cases where both an acute stroke and stroke sequelae are present, the acute episode takes precedence.

ICD-10 codes used:

I60 – Subarachnoid hemorrhage

I61 – Intracerebral hemorrhage

I62 – Other nontraumatic intracranial hemorrhages

I63 – Cerebral infarction

I64 – Stroke, not specified as hemorrhage or infarction

**Acute Heart Failure (MCV_ICA_AIG)**

This category includes patients hospitalized in short-stay medical and surgical units (MCO) during year n for heart failure, identified through the primary diagnosis (PD) of one or more hospital stays (RUMs), or for an acute complication of heart failure (e.g., hypertensive heart disease with heart failure, hypertensive heart and renal disease with or without heart failure, congestive hepatopathy, or acute pulmonary edema), with heart failure mentioned as an associated diagnosis (AD) or related diagnosis (RD).

In cases where both acute and chronic heart failure are coded, the acute episode takes precedence.

ICD-10 codes used (from PMSI):

Heart failure (primary diagnosis):

I50 – Heart failure

I11.0 – Hypertensive heart disease with (congestive) heart failure

I13.0 – Hypertensive heart and renal disease with heart failure

I13.2 – Hypertensive heart and renal disease with both (congestive) heart and renal failure

I13.9 – Hypertensive heart and renal disease, unspecified

K76.1 – Chronic passive congestion of the liver

J81 – Pulmonary edema

**Acute Pulmonary Embolism (MCV_EPU_IND)**

This group includes patients hospitalized during year n for pulmonary embolism, identified either by a primary diagnosis (PD) of pulmonary embolism in at least one hospital stay (RUM), or by the presence of a specific CCAM procedure performed during a hospital stay, in outpatient care, or in the private sector during the same year.

ICD-10 code used:

I26 – Pulmonary embolism

CCAM procedure codes used:

– Intraluminal dilation of the pulmonary artery: DFAF001, DFAF002, DFAF003, DFAF004

– Pulmonary artery clearance: DFFA001, DFFA002, DFFA003, DFNF001, DFNF002

**Supplementary Text 2. Expenditure categories**

The expenditure categories are from the HCEM and are defined by Rachas et al. in their article "The Economic Burden of Disease in France"[2].

| Group of Expenditure | Expenditure Category | Source of Data |
|---|---|---|
| Ambulatory care | General practitioners | Individual SNDS* reimbursement data |
| | Specialists | |
| | Dental care | |
| | Nursing care | |
| | Physiotherapists | |
| | Other paramedical care | |
| | Drugs | |
| | Medical devices and associated care | |
| | Laboratory tests | |
| | Other ambulatory care | |
| | Midwifery | |
| Hospital care | Transportation | Hospital discharge database |
| | Public hospital outpatient care | |
| | Short-stay hospitalization (DRG*) | |
| | Drugs and medical devices outside DRG* | |
| | Hospitalization in rehabilitation care | |
| | Hospitalization in psychiatry | |
| | Hospital at home | |
| Cash benefits | Maternity leave | Individual SNDS* reimbursement data |
| | Disability pension | |
| | Sick leave | |

*DRG: Diagnosis-related group

*SNDS: Système National des données de santé (French national health database)

**Supplementary Text 3**

To assess whether changes in healthcare reimbursements during the pandemic period were specific to patients with ACVD, we conducted a supplementary analysis comparing reimbursement disruptions between the ACVD group and a matched control group drawn from the general population, excluding patients with ACVD.

The control group was created using exact matching on age, sex, year of expenditure, COVID-19 status, and comorbidity level, generating 2,474 strata across both the ACVD and general population groups. To account for unequal numbers of individuals per stratum, we applied post-stratification weighting (weight = n_ACVD_stratum / n_control_stratum), resulting in a control group with seemingly identical size and structure to the ACVD cohort. Instead of matching each ACVD patient with a single control, we applied this weighting approach to leverage the full information available in the general population while reducing variance[3]. This process yielded a control group comprising 3,899,712 patient-years, identical in structure to the ACVD group. On average, annual healthcare expenditure in the control group was €10,963 per patient, compared to €21,999 for the ACVD population.

Similar to the ACVD population, an overall excess expenditure was estimated since the onset of the pandemic (2020–2023), totaling €1,969,800,492 (€1,963,595,840; €1,975,445,854), representing a 10.9% increase compared to pre-pandemic trends. Specifically, excess expenditures of €140,304,793 (€137,483,373; €142,915,528) in 2020, €442,406,042 (€440,211,046; €445,090,193) in 2021, €615,699,480 (€613,094,124; €618,963,993) in 2022, and €771,390,177 (€768,706,312; €773,754,603) in 2023 were estimated, corresponding to annual increases from 3.2% to 16.8% relative to expected values.

As with the ACVD group, these excess expenditures were primarily driven by patients also hospitalized for COVID-19 during the year: 50,6% of the total excess expenditure was attributable to only 3.8% of the population who had been hospitalized for COVID-19.

The categories of expenditure most impacted in the control group were similar to those observed in the ACVD population (see Supplementary Figures 1–5). However, the control group experienced a higher overall impact. Between 2020 and 2023, the excess expenditure reached 10.9% in the control group, compared to 6.2% in ACVD patients, yielding a ratio of 1.8. A ratio above 1 was observed across the main expenditure categories (see Supplementary Table 1).
A notable exception was observed in hospitalizations in psychiatry, where ACVD patients experienced excess expenditures 1.4 times higher than those of the control group.

Taken together, these findings suggest that the impact of the pandemic on healthcare reimbursement was not specific to patients with ACVD. Indeed, although patients with ACVD experienced significant excess expenditures, their increase was 1.8 times lower than that observed in the control group. This may reflect the effect of protective measures or specific care adaptations implemented for patients with ACVD during the pandemic period.

**Supplementary Figure 1. Observed minus expected expenditure per category and COVID-19 status, control group, France, 2020–2023**

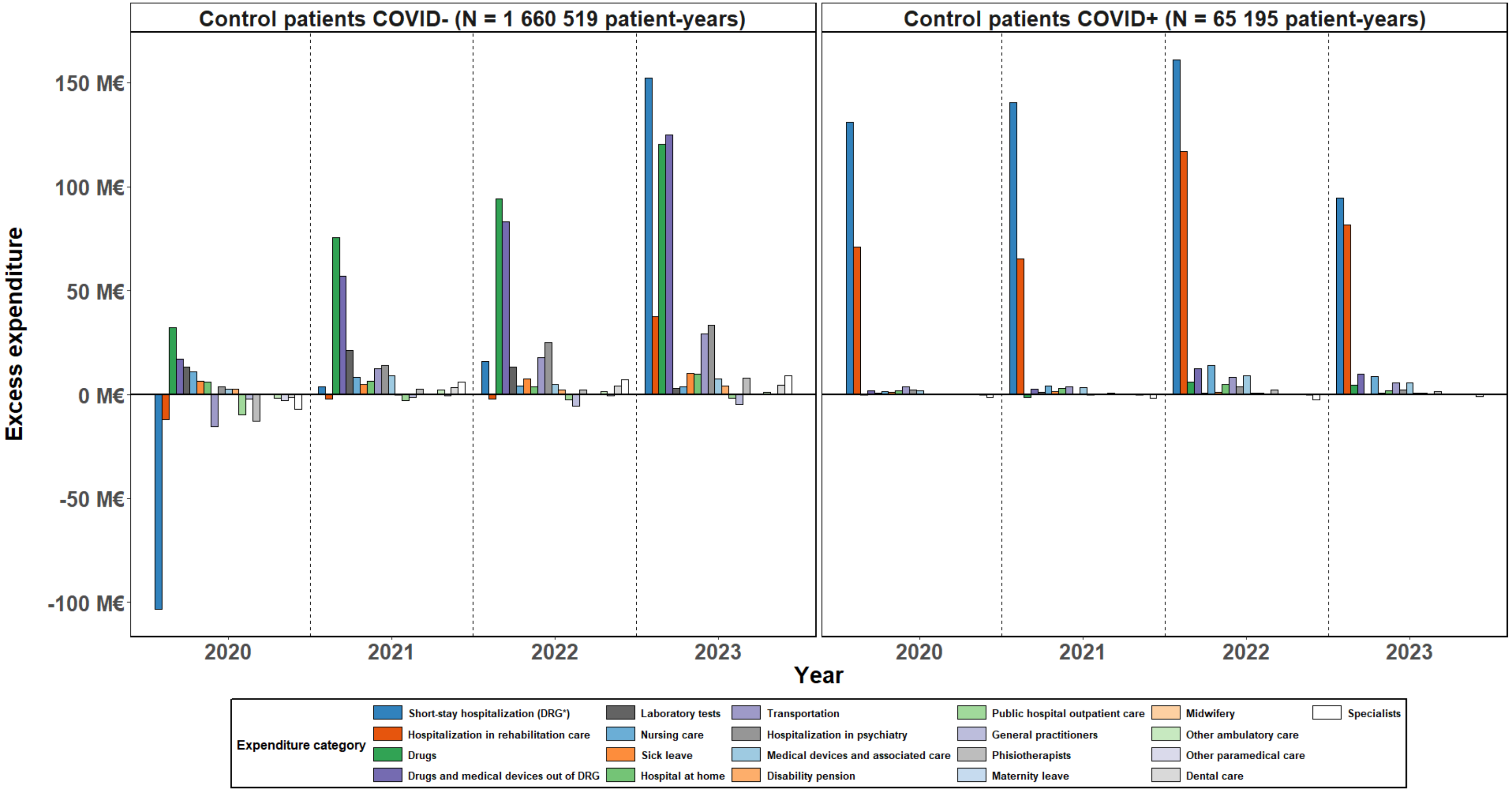


*DRG: diagnosis-related group; COVID-: Patients not hospitalized for COVID-19 reason within the year; COVID+: Patients hospitalized for COVID-19 reason within the year

Annual difference between observed and expected reimbursed amounts, by year, expenditure category, and COVID-19 status, for the control group. For example, in 2020, among patients of the control group hospitalized for COVID-19 (right panel), expenditures related to short-stay hospitalizations (dark blue bar) exceeded expected values (based on 2015–2019 trend) by more than €130 million.

**Supplementary Figure 2. Estimated observed minus expected expenditure by category, control group COVID+, France, 2020–2023**

| Expenditure category | 2020 (+216 M€) | 2021 (+223 M€) | 2022 (+340 M€) | 2023 (+218 M€) |
|---|---|---|---|---|
| Specialists | -1.5 M€ [-1.6; -1.5] (-23%) | -2 M€ [-2; -1.9] (-29%) | -2.6 M€ [-2.7; -2.6] (-27%) | -1.1 M€ [-1.1; -1] (-19%) |
| Dental care | -0.2 M€ [-0.2; -0.2] (-32%) | -0.2 M€ [-0.2; -0.2] (-22%) | -0.2 M€ [-0.2; -0.2] (-20%) | -59985 € [-61671; -58349] (-10%) |
| Other ambulatory care | -86352 € [-88025; -84522] (-69%) | -12286 € [-13197; -11380] (-10%) | +844 € [-598; 2327] (+1%) | -8889 € [-9766; -8113] (-12%) |
| Other paramedical care | -0.2 M€ [-0.2; -0.2] (-25%) | -0.3 M€ [-0.3; -0.3] (-35%) | -0.1 M€ [-0.2; -0.1] (-14%) | -1377 € [-5001; 2681] (0%) |
| Midwifery | -452 € [-746; -94] (-7%) | +1892 € [1337; 2700] (+26%) | -725 € [-1703; 263] (-9%) | -92 € [-702; 597] (-2%) |
| Maternity leave | +5780 € [2815; 8460] (+35%) | +16554 € [12389; 22064] (+82%) | +15779 € [8512; 24384] (+90%) | +1973 € [-1479; 6904] (+30%) |
| Laboratory tests | +0.9 M€ [0.9; 0.9] (+34%) | +1 M€ [1; 1] (+40%) | +0.9 M€ [0.9; 0.9] (+26%) | +0.2 M€ [0.2; 0.2] (+10%) |
| General practitioners | +0.2 M€ [0.2; 0.2] (+5%) | -0.1 M€ [-0.2; -0.1] (-4%) | +11173 € [6774; 16297] (+0%) | +0.2 M€ [0.2; 0.2] (+7%) |
| Public hospital outpatient care | +0.2 M€ [0.2; 0.3] (+7%) | +0.1 M€ [0.1; 0.1] (+3%) | +0.8 M€ [0.8; 0.8] (+17%) | +0.7 M€ [0.7; 0.7] (+23%) |
| Sick leave | +1 M€ [1; 1.1] (+68%) | +1.6 M€ [1.5; 1.6] (+91%) | +1.3 M€ [1.2; 1.4] (+66%) | +0.7 M€ [0.7; 0.8] (+67%) |
| Disability pension | -0.1 M€ [-0.1; -0.1] (-6%) | -0.3 M€ [-0.3; -0.3] (-13%) | +0.7 M€ [0.7; 0.8] (+28%) | +0.7 M€ [0.7; 0.8] (+50%) |
| Phisiotherapists | +0.2 M€ [0.1; 0.2] (+3%) | +0.8 M€ [0.7; 0.8] (+15%) | +2.1 M€ [2.1; 2.2] (+30%) | +1.7 M€ [1.7; 1.7] (+40%) |
| Hospital at home | +1.9 M€ [1.9; 2] (+54%) | +3 M€ [2.9; 3] (+80%) | +4.8 M€ [4.7; 4.9] (+87%) | +2 M€ [2; 2.1] (+60%) |
| Hospitalization in psychiatry | +2.1 M€ [2.1; 2.2] (+52%) | +0.5 M€ [0.5; 0.6] (+12%) | +3.7 M€ [3.5; 3.8] (+70%) | +2.2 M€ [2.1; 2.3] (+74%) |
| Drugs | -0.4 M€ [-0.5; -0.3] (-1%) | -1.6 M€ [-1.6; -1.5] (-6%) | +6.2 M€ [6; 6.3] (+16%) | +4.6 M€ [4.5; 4.7] (+20%) |
| Medical devices and associated care | +2.1 M€ [2; 2.1] (+22%) | +3.5 M€ [3.4; 3.5] (+36%) | +9.3 M€ [9.1; 9.4] (+69%) | +5.7 M€ [5.6; 5.8] (+72%) |
| Transportation | +4 M€ [3.9; 4.1] (+43%) | +4 M€ [3.9; 4.1] (+43%) | +8.2 M€ [8.1; 8.3] (+64%) | +5.7 M€ [5.6; 5.8] (+77%) |
| Nursing care | +1.6 M€ [1.5; 1.6] (+8%) | +4.2 M€ [4.1; 4.2] (+20%) | +14.2 M€ [14; 14.4] (+48%) | +8.8 M€ [8.7; 9] (+50%) |
| Drugs and medical devices out of DRG | +2 M€ [1.9; 2.1] (+18%) | +2.8 M€ [2.7; 2.9] (+24%) | +12.5 M€ [12.1; 12.8] (+74%) | +9.9 M€ [9.6; 10.3] (+97%) |
| Hospitalization in rehabilitation care | +71.2 M€ [69.9; 72.3] (+404%) | +65.5 M€ [64.5; 66.4] (+388%) | +116.9 M€ [115.5; 118.5] (+493%) | +81.8 M€ [80.6; 83.2] (+596%) |
| Short-stay hospitalization (DRG*) | +131 M€ [128.9; 133.4] (+190%) | +140.6 M€ [138.7; 142.8] (+209%) | +160.9 M€ [158.9; 163.2] (+173%) | +94.5 M€ [92.7; 96.3] (+177%) |

Year

*DRG: diagnosis-related group.

For each year and expenditure category, the color gradient reflects the total estimated excess expenditure among control group patients with at least one COVID-19-related hospitalization during the year. Excess expenditure is defined as the difference between the observed amount and the expected amount based on pre-pandemic trends (2015–2019) over the 2020–2023 period. The crude amount is reported (with 95% confidence interval), as well as the relative percentage compared to expected values. For example, in 2020, observed expenditures for short-stay hospitalization were €131 million above those expected based on pre-pandemic years. This excess expenditure corresponded to a rise of 190% compared to expected expenditures.

**Supplementary Figure 3. Estimated observed minus expected expenditure by category and by patient, control group COVID+, France, 2020–2023**

| Expenditure category | 2020 (+13704 € / patient) | 2021 (+13856 € / patient) | 2022 (+16043 € / patient) | 2023 (+17915 € / patient) |
|---|---|---|---|---|
| Specialists | -97 € [-98.8; -94.8] (-23%) | -123 € [-126; -120.7] (-29%) | -124 € [-126.7; -121.7] (-27%) | -88 € [-91.6; -84.9] (-19%) |
| Dental care | -15 € [-14.9; -14.4] (-32%) | -10 € [-10.6; -10.2] (-22%) | -9 € [-9.5; -9.2] (-20%) | -5 € [-5.1; -4.8] (-10%) |
| Other ambulatory care | -5 € [-5.6; -5.4] (-69%) | -1 € [-0.8; -0.7] (-10%) | +0 € [0; 0.1] (+1%) | -1 € [-0.8; -0.7] (-12%) |
| Other paramedical care | -11 € [-11.3; -10.9] (-25%) | -16 € [-16.2; -15.6] (-35%) | -7 € [-7.1; -6.7] (-14%) | 0 € [-0.4; 0.2] (0%) |
| Midwifery | 0 € [0; 0] (-7%) | +0 € [0.1; 0.2] (+26%) | 0 € [-0.1; 0] (-9%) | 0 € [-0.1; 0] (-2%) |
| Maternity leave | +0 € [0.2; 0.5] (+35%) | +1 € [0.8; 1.4] (+82%) | +1 € [0.4; 1.2] (+90%) | +0 € [-0.1; 0.6] (+30%) |
| Laboratory tests | +55 € [54.5; 56.4] (+34%) | +63 € [62.2; 64.6] (+40%) | +42 € [41.4; 42.7] (+26%) | +16 € [15.8; 16.7] (+10%) |
| General practitioners | +11 € [10.2; 11.1] (+5%) | -9 € [-9.6; -8.5] (-4%) | +1 € [0.3; 0.8] (+0%) | +18 € [17.1; 18] (+7%) |
| Public hospital outpatient care | +15 € [14.6; 16.3] (+7%) | +7 € [5.9; 7.3] (+3%) | +39 € [38.1; 39.7] (+17%) | +55 € [53.7; 56.7] (+23%) |
| Sick leave | +65 € [61.9; 68.2] (+68%) | +97 € [93.2; 102.2] (+91%) | +61 € [58.7; 65.4] (+66%) | +60 € [55.7; 64.7] (+67%) |
| Disability pension | -8 € [-8.9; -7.1] (-6%) | -19 € [-20.7; -18.2] (-13%) | +35 € [32.9; 38] (+28%) | +61 € [56.4; 66] (+50%) |
| Phisiotherapists | +10 € [8.7; 10.5] (+3%) | +48 € [46; 48.9] (+15%) | +101 € [99.2; 102.6] (+30%) | +139 € [136.4; 141.4] (+40%) |
| Hospital at home | +123 € [120.7; 126.8] (+54%) | +185 € [180.9; 189.1] (+80%) | +228 € [223.5; 231.6] (+87%) | +166 € [161.8; 170.7] (+60%) |
| Hospitalization in psychiatry | +136 € [131.4; 141.5] (+52%) | +33 € [28.1; 36.6] (+12%) | +175 € [167.2; 180] (+70%) | +182 € [173.2; 191] (+74%) |
| Drugs | -26 € [-32.4; -18] (-1%) | -97 € [-101.8; -92] (-6%) | +292 € [284.9; 297.4] (+16%) | +376 € [365.9; 388.8] (+20%) |
| Medical devices and associated care | +132 € [130; 135.2] (+22%) | +217 € [213.5; 220.2] (+36%) | +438 € [431.4; 444.5] (+69%) | +466 € [457.9; 475.2] (+72%) |
| Transportation | +252 € [246.3; 257.6] (+43%) | +247 € [242.7; 251.7] (+43%) | +388 € [381.4; 393.7] (+64%) | +468 € [459.1; 478.7] (+77%) |
| Nursing care | +99 € [93.9; 104.3] (+8%) | +258 € [251.7; 264] (+20%) | +670 € [660.3; 678.3] (+48%) | +723 € [710.9; 734.7] (+50%) |
| Drugs and medical devices out of DRG | +125 € [118.6; 130.2] (+18%) | +173 € [167.1; 182.1] (+24%) | +589 € [573.6; 603.5] (+74%) | +813 € [786.5; 842.2] (+97%) |
| Hospitalization in rehabilitation care | +4519 € [4436.7; 4592.2] (+404%) | +4068 € [4006.7; 4125.8] (+388%) | +5523 € [5458.5; 5599.8] (+493%) | +6712 € [6608.8; 6824.7] (+596%) |
| Short-stay hospitalization (DRG*) | +8322 € [8186.2; 8474] (+190%) | +8735 € [8619.4; 8873] (+209%) | +7603 € [7507.6; 7711.9] (+173%) | +7752 € [7600.9; 7898.7] (+177%) |

Year

*DRG: diagnosis-related group.

For each year and expenditure category, the color gradient reflects the estimated excess expenditure by patient among control group patients with at least one COVID-19-related hospitalization during the year. The average amount is reported (with 95% confidence interval), as well as the relative percentage compared to expected values. For example, in 2020, observed expenditures for short-stay hospitalization per patient were €8322 above those expected based on pre-pandemic years. This excess expenditure corresponded to a rise of 190% compared to expected expenditures.

**Supplementary Figure 4. Estimated observed minus expected expenditure by category, control group COVID-, France, 2020–2023**

| Expenditure category | 2020 (-75 M€) | 2021 (+219 M€) | 2022 (+276M€) | 2023 (+553M€) |
|---|---|---|---|---|
| General practitioners | -2.4 M€ [-2.4; -2.4] (-3%) | -1.6 M€ [-1.6; -1.6] (-2%) | -5.6 M€ [-5.6; -5.6] (-7%) | -5 M€ [-5; -4.9] (-6%) |
| Public hospital outpatient care | -10 M€ [-10; -10] (-13%) | -3 M€ [-3; -3] (-4%) | -2.5 M€ [-2.5; -2.5] (-3%) | -1.9 M€ [-1.9; -1.9] (-2%) |
| Midwifery | -20656 € [-20911; -20370] (-7%) | +5571 € [5201; 6051] (+2%) | +27795 € [27358; 28172] (+9%) | +52892 € [52439; 53363] (+18%) |
| Maternity leave | +2424 € [741; 4060] (+0%) | +65551 € [62190; 68456] (+5%) | +87958 € [84879; 91224] (+7%) | +70395 € [67296; 73708] (+6%) |
| Other paramedical care | -3.1 M€ [-3.1; -3] (-20%) | -0.8 M€ [-0.8; -0.8] (-5%) | -0.6 M€ [-0.6; -0.6] (-4%) | +0.2 M€ [0.2; 0.2] (+1%) |
| Other ambulatory care | -2.1 M€ [-2.1; -2.1] (-57%) | +2.4 M€ [2.4; 2.4] (+67%) | +1.6 M€ [1.6; 1.6] (+49%) | +1.2 M€ [1.2; 1.2] (+38%) |
| Laboratory tests | +13.1 M€ [13.1; 13.2] (+23%) | +21.4 M€ [21.4; 21.5] (+37%) | +13.3 M€ [13.3; 13.4] (+24%) | +3 M€ [2.9; 3] (+5%) |
| Nursing care | +11 M€ [11; 11] (+3%) | +8.5 M€ [8.5; 8.5] (+2%) | +4.1 M€ [4.1; 4.2] (+1%) | +3.7 M€ [3.6; 3.8] (+1%) |
| Disability pension | +2.5 M€ [2.4; 2.5] (+3%) | -0.4 M€ [-0.5; -0.4] (0%) | +2.4 M€ [2.3; 2.5] (+2%) | +4.2 M€ [4.1; 4.3] (+4%) |
| Dental care | -1.6 M€ [-1.6; -1.6] (-7%) | +3.3 M€ [3.3; 3.3] (+15%) | +4 M€ [4; 4] (+18%) | +4.7 M€ [4.7; 4.7] (+20%) |
| Medical devices and associated care | +2.5 M€ [2.5; 2.5] (+1%) | +9.1 M€ [9; 9.1] (+4%) | +5.1 M€ [5.1; 5.1] (+2%) | +7.5 M€ [7.5; 7.6] (+3%) |
| Phisiotherapists | -12.9 M€ [-12.9; -12.8] (-12%) | +2.6 M€ [2.6; 2.6] (+2%) | +2.2 M€ [2.2; 2.2] (+2%) | +8.1 M€ [8.1; 8.2] (+7%) |
| Specialists | -7.3 M€ [-7.3; -7.2] (-5%) | +6.1 M€ [6; 6.1] (+4%) | +7 M€ [7; 7.1] (+4%) | +9.2 M€ [9.2; 9.3] (+5%) |
| Hospital at home | +6 M€ [6; 6] (+9%) | +6.5 M€ [6.5; 6.5] (+9%) | +3.6 M€ [3.6; 3.7] (+5%) | +9.8 M€ [9.7; 9.8] (+12%) |
| Sick leave | +6.5 M€ [6.5; 6.5] (+9%) | +5 M€ [4.9; 5] (+7%) | +7.5 M€ [7.4; 7.6] (+10%) | +10.3 M€ [10.2; 10.4] (+13%) |
| Transportation | -15.5 M€ [-15.5; -15.4] (-8%) | +12.5 M€ [12.5; 12.6] (+7%) | +17.8 M€ [17.7; 17.9] (+10%) | +29.2 M€ [29.1; 29.3] (+15%) |
| Hospitalization in psychiatry | +3.9 M€ [3.8; 3.9] (+3%) | +14.1 M€ [14; 14.2] (+10%) | +25.2 M€ [25.1; 25.4] (+18%) | +33.5 M€ [33.3; 33.7] (+24%) |
| Hospitalization in rehabilitation care | -12.3 M€ [-12.3; -12.2] (-4%) | -2.4 M€ [-2.5; -2.3] (-1%) | -2.2 M€ [-2.3; -2.1] (-1%) | +37.6 M€ [37.5; 37.7] (+12%) |
| Drugs | +32.2 M€ [32.1; 32.3] (+5%) | +75.6 M€ [75.3; 75.9] (+12%) | +94 M€ [93.7; 94.3] (+15%) | +120.5 M€ [120.1; 120.9] (+19%) |
| Drugs and medical devices out of DRG | +17.3 M€ [17.2; 17.4] (+8%) | +56.8 M€ [56.5; 57.1] (+22%) | +83.1 M€ [82.7; 83.5] (+32%) | +124.7 M€ [124.1; 125.4] (+43%) |
| Short-stay hospitalization (DRG*) | -103.4 M€ [-103.9; -103.1] (-8%) | +3.6 M€ [3.4; 3.8] (+0%) | +16 M€ [15.7; 16.3] (+1%) | +152.3 M€ [151.6; 153] (+11%) |

Year

*DRG: diagnosis-related group.

For each year and expenditure category, the color gradient reflects the total estimated excess expenditure among control group patients without COVID-19-related hospitalization during the year. The crude amount is reported (with 95% confidence interval), as well as the relative percentage compared to expected values. For example, in 2023, observed expenditures for short-stay hospitalization were €152.3 million above those expected based on pre-pandemic years. This excess expenditure corresponded to a rise of 11% compared to expected expenditures.

**Supplementary Figure 5. Estimated observed minus expected expenditure by category and by patient, control group COVID-, France, 2020–2023**

| Expenditure category | 2020 (-184 € / patient) | 2021 (+517 € / patient) | 2022 (+675€ / patient) | 2023 (+1326€ / patient) |
|---|---|---|---|---|
| General practitioners | -6 € [-5.8; -5.7] (-3%) | -4 € [-3.8; -3.8] (-2%) | -14 € [-13.8; -13.7] (-7%) | -12 € [-12; -11.9] (-6%) |
| Public hospital outpatient care | -24 € [-24.5; -24.3] (-13%) | -7 € [-7.2; -7.1] (-4%) | -6 € [-6.2; -6.2] (-3%) | -4 € [-4.5; -4.4] (-2%) |
| Midwifery | 0 € [-0.1; 0] (-7%) | +0 € [0; 0] (+2%) | +0 € [0.1; 0.1] (+9%) | +0 € [0.1; 0.1] (+18%) |
| Maternity leave | +0 € [0; 0] (+0%) | +0 € [0.1; 0.2] (+5%) | +0 € [0.2; 0.2] (+7%) | +0 € [0.2; 0.2] (+6%) |
| Other paramedical care | -7 € [-7.5; -7.4] (-20%) | -2 € [-1.9; -1.8] (-5%) | -1 € [-1.4; -1.4] (-4%) | +1 € [0.5; 0.5] (+1%) |
| Other ambulatory care | -5 € [-5.1; -5] (-57%) | +6 € [5.6; 5.6] (+67%) | +4 € [3.9; 3.9] (+49%) | +3 € [2.9; 2.9] (+38%) |
| Laboratory tests | +32 € [31.9; 32.1] (+23%) | +51 € [50.3; 50.6] (+37%) | +33 € [32.5; 32.7] (+24%) | +7 € [7.1; 7.1] (+5%) |
| Nursing care | +27 € [26.7; 26.9] (+3%) | +20 € [20; 20.1] (+2%) | +10 € [9.9; 10.2] (+1%) | +9 € [8.7; 9.1] (+1%) |
| Disability pension | +6 € [6; 6.1] (+3%) | -1 € [-1.1; -0.9] (0%) | +6 € [5.7; 6] (+2%) | +10 € [9.8; 10.2] (+4%) |
| Dental care | -4 € [-3.8; -3.8] (-7%) | +8 € [7.8; 7.9] (+15%) | +10 € [9.8; 9.8] (+18%) | +11 € [11.2; 11.2] (+20%) |
| Medical devices and associated care | +6 € [6; 6.1] (+1%) | +21 € [21.3; 21.5] (+4%) | +12 € [12.4; 12.5] (+2%) | +18 € [18; 18.2] (+3%) |
| Phisiotherapists | -31 € [-31.5; -31.3] (-12%) | +6 € [6.1; 6.2] (+2%) | +5 € [5.4; 5.4] (+2%) | +20 € [19.4; 19.6] (+7%) |
| Specialists | -18 € [-17.8; -17.7] (-5%) | +14 € [14.2; 14.4] (+4%) | +17 € [17.1; 17.3] (+4%) | +22 € [22; 22.2] (+5%) |
| Hospital at home | +15 € [14.6; 14.7] (+9%) | +15 € [15.2; 15.4] (+9%) | +9 € [8.8; 9] (+5%) | +23 € [23.3; 23.5] (+12%) |
| Sick leave | +16 € [15.8; 16] (+9%) | +12 € [11.6; 11.8] (+7%) | +18 € [18.2; 18.5] (+10%) | +25 € [24.5; 24.8] (+13%) |
| Transportation | -38 € [-37.9; -37.6] (-8%) | +29 € [29.3; 29.6] (+7%) | +43 € [43.2; 43.6] (+10%) | +70 € [69.8; 70.3] (+15%) |
| Hospitalization in psychiatry | +9 € [9.4; 9.6] (+3%) | +33 € [33; 33.4] (+10%) | +62 € [61.3; 62] (+18%) | +80 € [79.8; 80.7] (+24%) |
| Hospitalization in rehabilitation care | -30 € [-30.1; -29.9] (-4%) | -6 € [-5.8; -5.5] (-1%) | -5 € [-5.5; -5.2] (-1%) | +90 € [89.8; 90.4] (+12%) |
| Drugs | +78 € [78.2; 78.7] (+5%) | +178 € [177.5; 178.7] (+12%) | +230 € [229.2; 230.7] (+15%) | +289 € [288; 289.8] (+19%) |
| Drugs and medical devices out of DRG | +42 € [41.8; 42.4] (+8%) | +134 € [133.1; 134.6] (+22%) | +203 € [202.2; 204.3] (+32%) | +299 € [297.6; 300.6] (+43%) |
| Short-stay hospitalization (DRG*) | -252 € [-253.3; -251.4] (-8%) | +9 € [8; 9.1] (+0%) | +39 € [38.5; 39.9] (+1%) | +365 € [363.5; 366.8] (+11%) |

Year

For each year and expenditure category, the color gradient reflects the estimated excess expenditure by patient among control group patients without COVID-19-related hospitalization during the year. The average amount is reported (with 95% confidence interval), as well as the relative percentage compared to expected values. For example, in 2023, observed expenditures for short-stay hospitalization per patient were €365 above those expected based on pre-pandemic years. This excess expenditure corresponded to a rise of 11% compared to expected expenditures.

**Supplementary Table 1: Excess expenditure ratio (control group versus ACVD group) by cost category and COVID-19 status**

| Expense category | Ratio (Total Patients) | Ratio (COVID+ Patients) | Ratio (COVID- Patients) |
|---|---|---|---|
| Total | 1.75 [1.74; 1.75] | 1.95 [1.94; 1.96] | 1.46 [1.46; 1.47] |
| Short-stay hospitalization | 2.45 [2.42; 2.47] | 3.35 [3.33; 3.37] | 0.62 [0.61; 0.63] |
| Hospitalization in rehabilitation care | 1.93 [1.91; 1.95] | 2.33 [2.32; 2.35] | 0.31 [0.31; 0.32] |
| Drugs | 0.96 [0.96; 0.96] | 0.69 [0.66; 0.72] | 0.97 [0.97; 0.97] |
| Transportation | 0.82 [0.81; 0.82] | 1.48 [1.47; 1.50] | 0.66 [0.66; 0.66] |
| Medical devices and associated care | 0.79 [0.78; 0.79] | 1.49 [1.48; 1.50] | 0.56 [0.56; 0.56] |
| Nursing care | 0.89 [0.88; 0.90] | 1.52 [1.51; 1.54] | 0.62 [0.62; 0.62] |
| Hospitalization in psychiatry | 0.70 [0.69; 0.71] | 0.95 [0.89; 1.02] | 0.69 [0.68; 0.69] |
| Laboratory tests | 1.11 [1.11; 1.11] | 1.38 [1.36; 1.39] | 1.10 [1.09; 1.10] |
| Drugs and medical devices out of DRG | 9.55 [9.35; 9.77] | 8.43 [7.61; 9.46] | 9.59 [9.37; 9.78] |
| Hospital at home | 1.23 [1.22; 1.24] | 1.74 [1.70; 1.79] | 1.07 [1.06; 1.08] |
| Sick leave | 4.59 [4.52; 4.67] | 6.60 [6.00; 7.50] | 4.32 [4.26; 4.37] |
| Physiotherapists | 0.39 [0.39; 0.40] | 1.36 [1.33; 1.38] | 0.01 [0.00; 0.02] |
| Dental care | 1.04 [1.04; 1.04] | 1.61 [1.58; 1.65] | 1.06 [1.06; 1.07] |
| Specialists | 1.28 [1.25; 1.30] | 1.61 [1.59; 1.64] | 1.50 [1.49; 1.52] |
| Other ambulatory care | 0.65 [0.64; 0.65] | -45.01 [-54.06; -17.46] | 0.67 [0.67; 0.68] |
| Disability pension | 3.62 [3.28; 3.98] | 0.62 [0.56; 0.69] | 9.70 [7.80; 12.81] |
| Maternity leave | 0.70 [0.62; 0.77] | 1.87 [0.88; 8.90] | 0.63 [0.56; 0.69] |
| Midwifery | -0.70 [-0.74; -0.66] | -0.03 [-0.06; 0.00] | -0.77 [-0.81; -0.74] |
| Other paramedical care | 2.46 [2.43; 2.49] | 0.97 [0.95; 0.99] | 3.01 [2.96; 3.05] |
| Public hospital outpatient care | 1.25 [1.24; 1.25] | 1.75 [1.71; 1.79] | 1.30 [1.30; 1.31] |
| General practitioners | 0.76 [0.76; 0.76] | -0.48 [-0.51; -0.45] | 0.79 [0.79; 0.80] |

For each expenditure category and COVID-19 status, the table reports the ratio of estimated excess expenditures (2020–2023) between the control group and the ACVD group. For example, Hospitalization in rehabilitation care expenditures were 466% higher than expected for patients in the control COVID+ group over the 2020–2023 period, compared to a 200% higher in the ACVD COVID+ group. This yields a ratio of 2.33, indicating that the increase in expenditures for this category was 2.33 times more pronounced in the control group than in the ACVD group.

### Supplementary Text 3. Variable selection and interaction terms

#### Variable selection

In order to guide the selection of variables to include in the model, exploratory analyses based on visual representations of the relationship between predictor variables and the outcome were conducted. Specifically, the annual mean reimbursed amount per patient was visualized according to various patient characteristics.

As illustrated in Supplementary Figure 6, which presents the mean reimbursed amount according to age group and sex, the corresponding relationship with age is non-linear: the average reimbursement increases up to the 55–59 age group, followed by a decrease in older age groups. Moreover, differences by sex appear to vary across age groups. Women had higher mean annual reimbursements than men up to the 65–69 age group, while thereafter, the pattern was reversed. These observations suggest that age should be treated as a categorical variable rather than a continuous one, and that an interaction term with sex should be considered since the effect of sex appears age-dependent.

**Supplementary Figure 6. Average annual amount reimbursed per patient-year with ACVD according to age class and sex, France, 2015–2019**

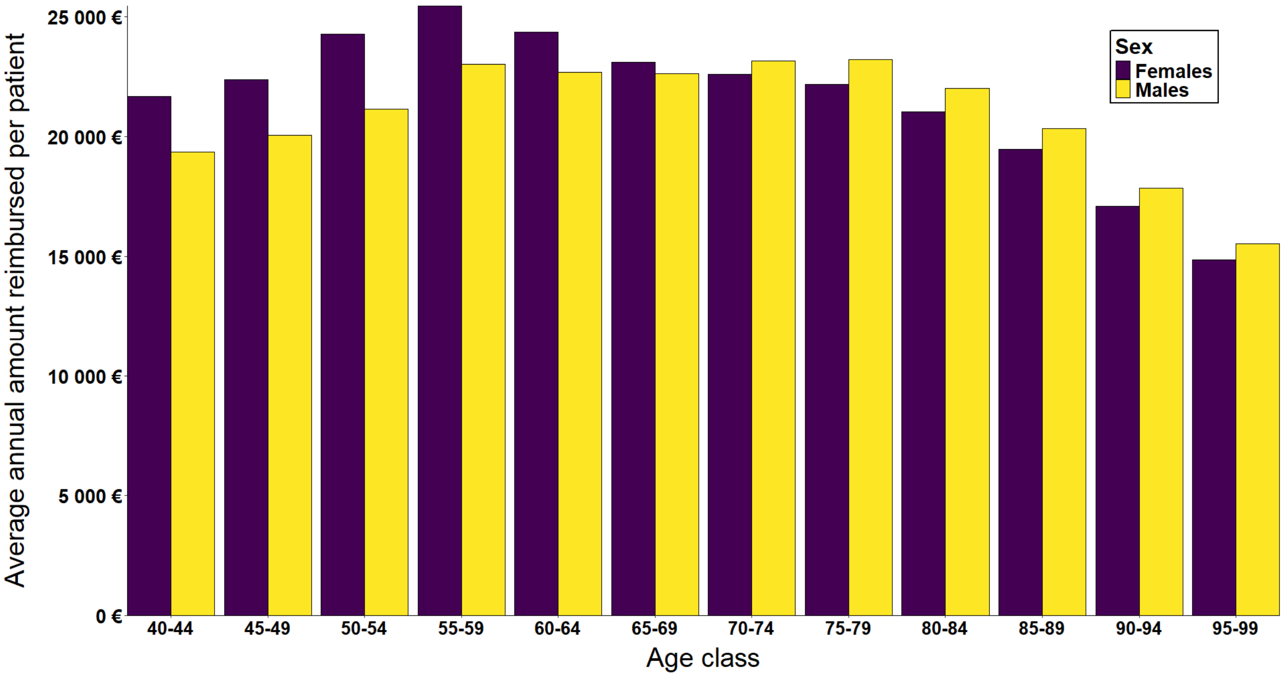


For each age class between 40-44 and 95-99 years, the bars represent the average annual amount reimbursed between 2015 and 2019 (purple for females and yellow for males). For example, the average annual amount reimbursed was about €22,000 for females between 40 and 44 years old (first purple bar) based on data from 2015 to 2019.

In order to take into account the potential role of socioeconomic variables, the relationship between healthcare reimbursements and social deprivation, measured with the French Deprivation Index[4], was also explored. Supplementary Figure 7 shows that the variations of the mean reimbursements across the quintiles of deprivation index were low, including a small difference between the least (Q1) and most (Q5) deprived groups. Therefore, this variable was not retained in the final model.

**Supplementary Figure 7. Average annual amount reimbursed per patient-year with ACVD according to social deprivation index, France, 2015–2019**

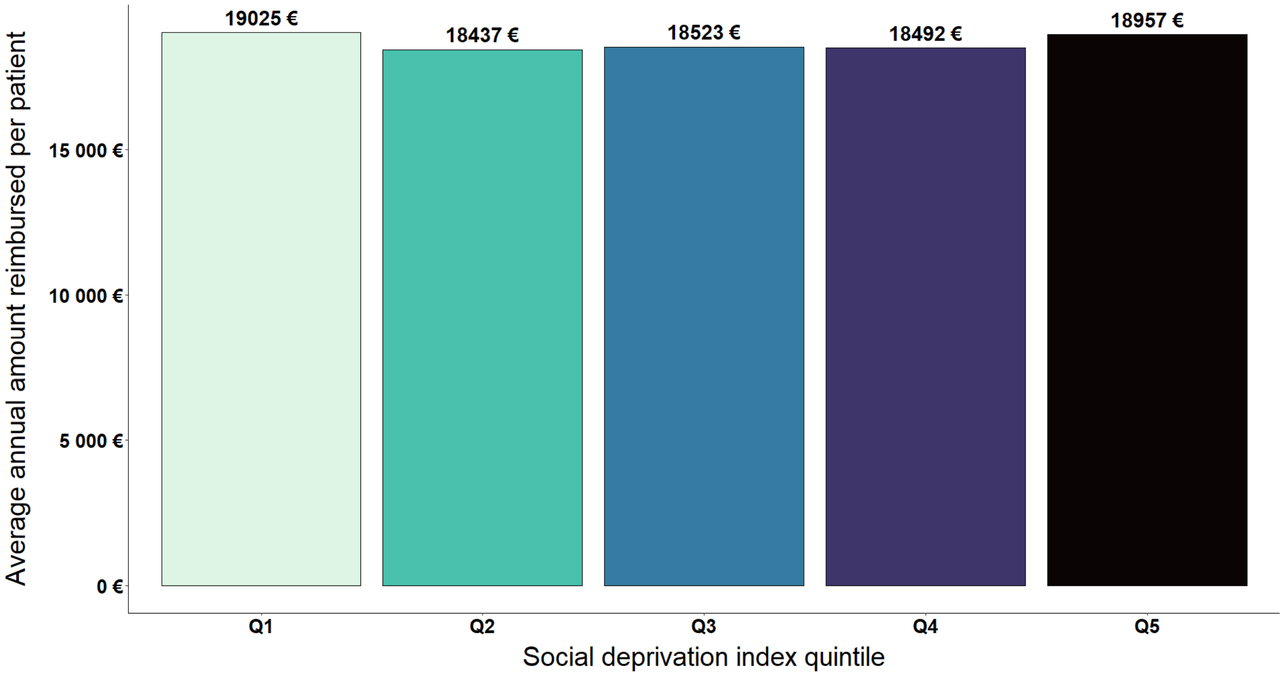


Each bar corresponds to the average annual amount reimbursed between 2015 and 2019 according to the social deprivation index, from Q1 (least deprived quintile of the French population) to Q5 (most deprived quintile of the French population).

In contrast, Supplementary Figure 8 shows a strong association between the Mortality-Related Morbidity Index[5] and the mean annual reimbursement. Therefore, this index was a variable included in the model.

**Supplementary Figure 8. Average annual amount reimbursed per patient-year with ACVD according to the comorbidity index, France, 2015–2019**

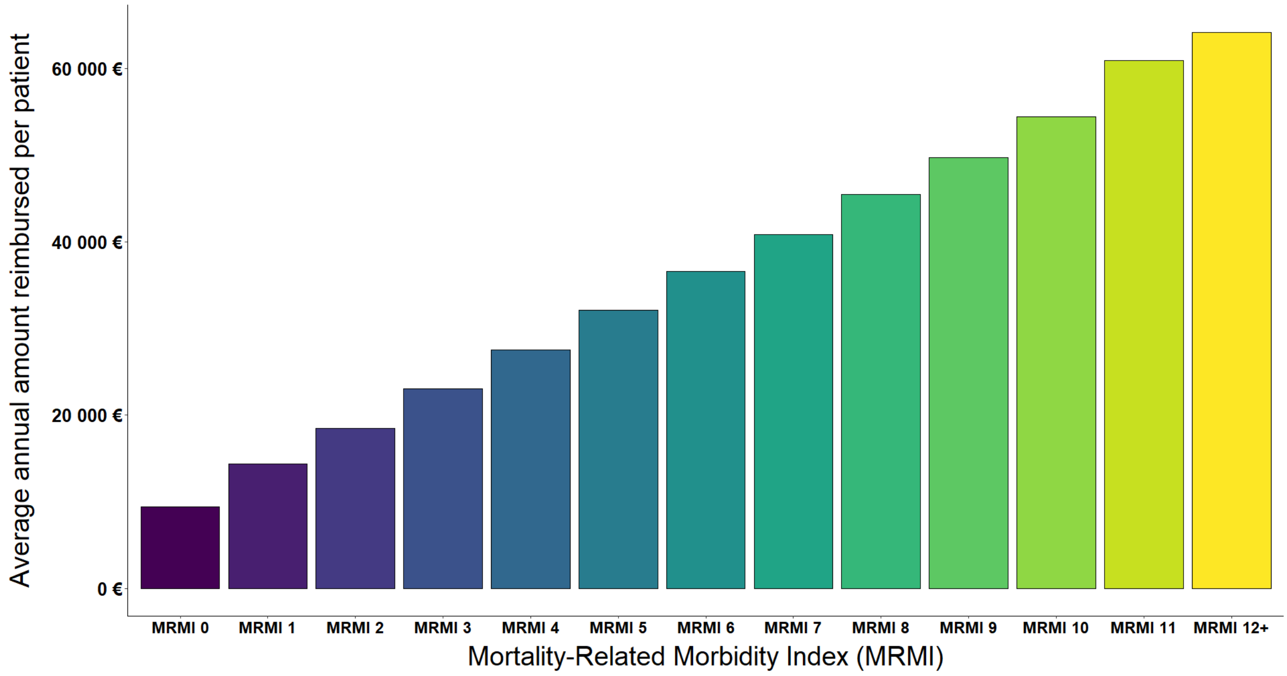


Each bar corresponds to the average annual amount reimbursed between 2015 and 2019 according to the comorbidity index, from 0 (no comorbidities) to 12+ (12 to 15 comorbidities).

### Interaction terms

To evaluate the model's ability to predict expected healthcare expenditures in a non-pandemic period, the model was trained on data from 2015 to 2018 and used to generate predictions for the year 2019. Since 2019 is preceding the pandemic period, and under the assumption that the model estimates expected non-pandemic expenditures, model performance was assessed by minimizing the difference between observed and predicted spending in 2019. Various combinations of selected predictor variables were tested, ranging from the simplest model with no interaction terms to the most complex specification including interactions between all four selected predictors (all interaction terms between 2 predictors, all interaction terms between 3 predictors, interaction between the four predictors).

As illustrated in Supplementary Figure 9, models with no interaction between predictors performed poorly, with prediction errors exceeding €800 million, more than 8% error. In contrast, models including interaction terms involving all combinations up to three predictors or combinations up to four predictors yielded predictions closely aligned with the actual observed 2019 expenditures, with errors under 0.1%. Notably, the "Fourth-order interactions" model predicted 2019 spending almost perfectly, with an error of only 0.5 million euros (-0.8; +1.8). Given that total observed spending for that year was 9,636 million euros, this corresponds to a relative error of just 0.01% (–0.01%; +0.02%).

**Supplementary Figure 9. Difference between observed and expected expenditures in 2019 based on data from 2015 to 2018, according to the model used**

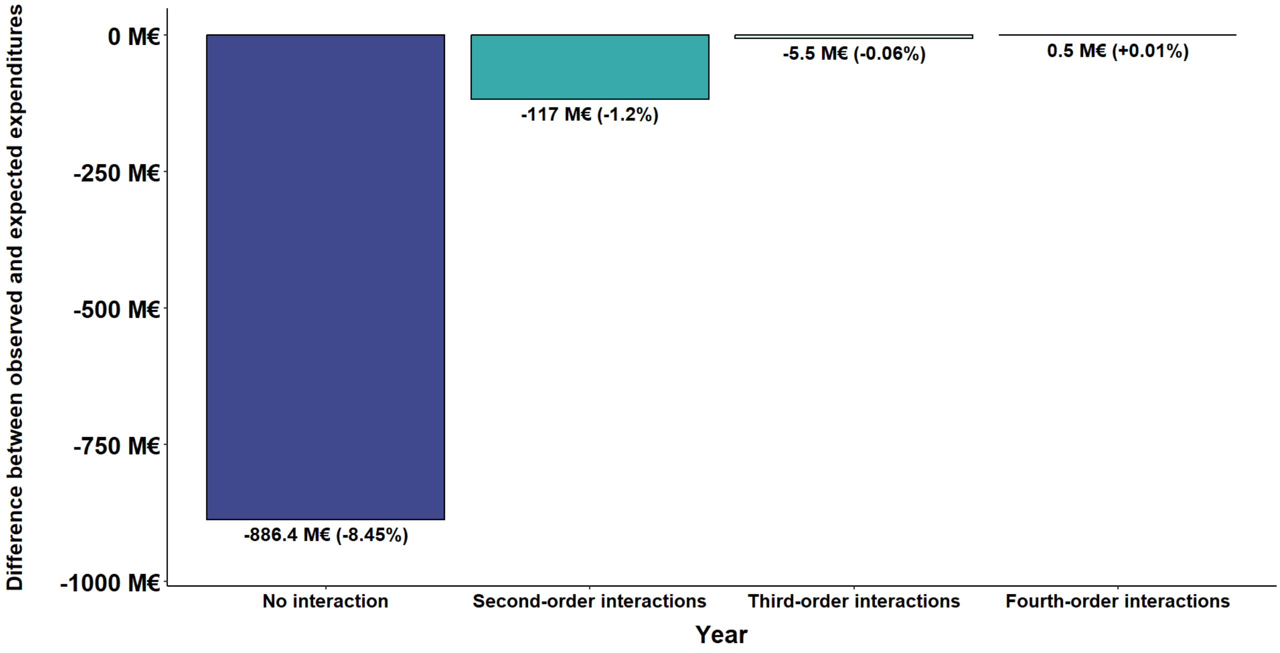


For each model, from no interaction between variables (No interaction) to interaction terms including all interaction terms up to the term combining all four variables (Fourth-order interactions), the bar represents the difference between expenditures observed in 2019 and those expected, expectations being based on years 2015 to 2018. This difference is expressed with crude (and relative values in parentheses). For example, an excess of €886.4 million in expenditures was estimated by the model without interaction for 2019 as compared to the expenditures actually observed this year (dark blue bar), with a corresponding overestimation of 8.45%.

To assess how the choice of model influenced the estimation of excess expenditures during the pandemic, the top two performing models were used to estimate excess spending from 2020 to 2023. As shown in Supplementary Figure 10, both models estimate an increase in healthcare expenditures during the pandemic period, including worsening excess over time. Given its superior predictive performance in 2019, the “Fourth-order interactions” model was selected as the primary model for analysis. Nevertheless, the findings remained consistent regardless of the model used.

**Supplementary Figure 10. Difference between observed and expected expenditures in the pandemic period (2020–2023) based on data from 2015 to 2019, according to the model used**

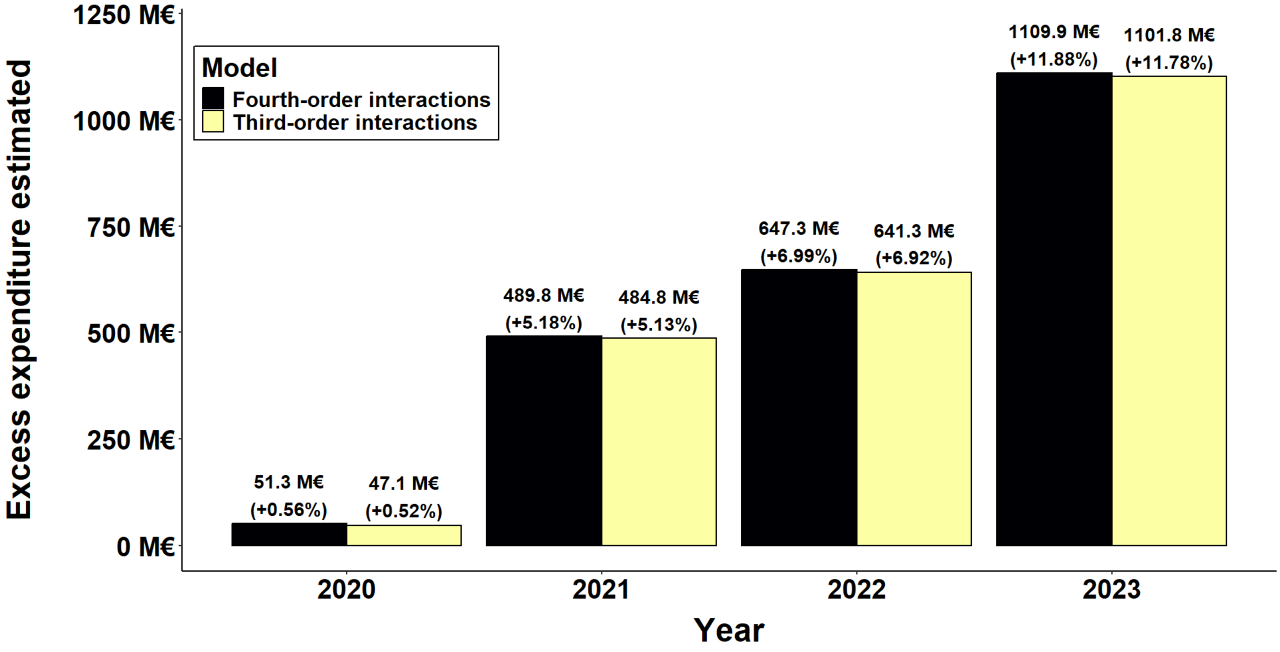


For two models, the bar represents the difference between expenditures observed in the pandemic period (2020 to 2023) and those expected based on the years 2015 to 2019. This difference is expressed with crude and relative values. For example, an estimation of €489.8 million excess expenditure was made in 2021 by the model with interactions between all variables (black bar). This excess expenditure corresponds to a rise of 5.18% compared to expected values based on the trends of years 2015 to 2019.

**Supplementary Figure 11. Estimated observed minus expected expenditure by category, ACVD COVID+, France, 2020–2023**

| Expenditure category | 2020 (+203 M€) | 2021 (+204 M€) | 2022 (+318 M€) | 2023 (+214 M€) |
|---|---|---|---|---|
| Specialists | -0.9 M€ [-0.9; -0.8] (-14%) | -1.2 M€ [-1.2; -1.2] (-19%) | -1.4 M€ [-1.5; -1.4] (-17%) | -0.5 M€ [-0.5; -0.5] (-10%) |
| Other paramedical care | -0.2 M€ [-0.2; -0.2] (-22%) | -0.3 M€ [-0.3; -0.2] (-32%) | -0.1 M€ [-0.2; -0.1] (-13%) | -67917 € [-73595; -61153] (-11%) |
| General practitioners | -13617 € [-20016; -8229] (0%) | -0.3 M€ [-0.3; -0.3] (-6%) | -0.3 M€ [-0.3; -0.2] (-4%) | -57137 € [-64622; -50305] (-2%) |
| Midwifery | -1496 € [-2483; -321] (-19%) | -3744 € [-4697; -2739] (-47%) | -4067 € [-5947; -2310] (-41%) | -2103 € [-3352; -201] (-41%) |
| Dental care | -0.1 M€ [-0.1; -0.1] (-25%) | -0.1 M€ [-0.1; -0.1] (-18%) | -70750 € [-74504; -67460] (-9%) | +4252 € [1140; 8198] (+1%) |
| Maternity leave | +20546 € [6783; 37708] (+72%) | -2775 € [-13329; 10492] (-8%) | +7872 € [-11887; 28898] (+30%) | +7067 € [-3685; 21567] (+88%) |
| Other ambulatory care | -38392 € [-40635; -36479] (-42%) | -2572 € [-3680; -1558] (-3%) | +22668 € [20945; 24540] (+23%) | +19986 € [18337; 21578] (+40%) |
| Laboratory tests | +0.7 M€ [0.7; 0.7] (+22%) | +1 M€ [1; 1] (+31%) | +0.8 M€ [0.8; 0.9] (+20%) | +0.1 M€ [0.1; 0.1] (+5%) |
| Disability pension | +0.2 M€ [0.2; 0.2] (+12%) | +0.3 M€ [0.3; 0.4] (+19%) | +0.5 M€ [0.5; 0.6] (+25%) | +0.3 M€ [0.2; 0.3] (+23%) |
| Public hospital outpatient care | +18453 € [5486; 29638] (+0%) | +0.1 M€ [0.1; 0.1] (+3%) | +0.6 M€ [0.6; 0.7] (+11%) | +0.5 M€ [0.5; 0.5] (+14%) |
| Sick leave | +0.3 M€ [0.3; 0.4] (+10%) | -12891 € [-75028; 52453] (0%) | +0.5 M€ [0.4; 0.6] (+13%) | +0.6 M€ [0.5; 0.7] (+29%) |
| Hospitalization in psychiatry | +2.2 M€ [2; 2.4] (+86%) | +1.2 M€ [1.1; 1.4] (+47%) | +1.1 M€ [1; 1.2] (+34%) | +1 M€ [0.8; 1.2] (+53%) |
| Hospital at home | +1.3 M€ [1.2; 1.4] (+26%) | +3.7 M€ [3.6; 3.8] (+72%) | +3.2 M€ [3; 3.4] (+42%) | +1 M€ [1; 1.1] (+23%) |
| Phisiotherapists | -89365 € [-103007; -77418] (-2%) | +0.9 M€ [0.9; 0.9] (+19%) | +1.5 M€ [1.5; 1.5] (+21%) | +1.1 M€ [1.1; 1.1] (+27%) |
| Drugs and medical devices out of DRG | -1.2 M€ [-1.3; -1.1] (-7%) | -1.2 M€ [-1.4; -1] (-7%) | +3.7 M€ [3.3; 4.1] (+14%) | +3.8 M€ [3.5; 4.1] (+24%) |
| Drugs | +0.7 M€ [0.6; 0.8] (+2%) | +1.2 M€ [1.1; 1.3] (+4%) | +6.9 M€ [6.6; 7.3] (+18%) | +3.8 M€ [3.7; 4] (+17%) |
| Medical devices and associated care | +2.8 M€ [2.8; 2.9] (+21%) | +4.4 M€ [4.3; 4.4] (+32%) | +7.6 M€ [7.4; 7.7] (+41%) | +4.3 M€ [4.2; 4.4] (+39%) |
| Transportation | +2.4 M€ [2.3; 2.5] (+19%) | +4 M€ [3.9; 4.1] (+33%) | +7.7 M€ [7.6; 7.9] (+47%) | +5.1 M€ [5; 5.2] (+54%) |
| Nursing care | +3 M€ [3; 3.1] (+13%) | +4.4 M€ [4.3; 4.5] (+18%) | +9.5 M€ [9.3; 9.6] (+27%) | +5.2 M€ [5.1; 5.3] (+25%) |
| Short-stay hospitalization (DRG*) | +106.4 M€ [104.5; 108.2] (+56%) | +112 M€ [109.8; 113.9] (+59%) | +131.3 M€ [128.5; 133.7] (+51%) | +87.4 M€ [85.6; 89.4] (+60%) |
| Hospitalization in rehabilitation care | +85 M€ [83.4; 86.4] (+173%) | +73.9 M€ [72.6; 75.1] (+151%) | +144.9 M€ [143.1; 146.7] (+219%) | +100.6 M€ [98.9; 102.9] (+264%) |

Year

*DRG: diagnosis-related group.

For each year and expenditure category, the color gradient reflects the total estimated excess expenditure among ACVD patients with at least one COVID-19-related hospitalization during the year. Excess expenditure is defined as the difference between the observed amount and the expected amount based on pre-pandemic trends (2015–2019) over the 2020–2023 period. The crude amount is reported (with 95% confidence interval), as well as the relative percentage compared to expected values. For example, in 2020, observed expenditures for short-stay hospitalization were €106.4 million above those expected based on pre-pandemic years. This excess expenditure corresponded to a rise of 56% compared to expected expenditures.

**Supplementary Figure 12. Estimated observed minus expected expenditure by category and by patient, ACVD COVID+, France, 2020–2023**

| Expenditure category | 2020 (+12869 € / patient) | 2021 (+12678 € / patient) | 2022 (+15028 € / patient) | 2023 (+17570 € / patient) |
|---|---|---|---|---|
| Specialists | -55 € [-56.2; -53] (-14%) | -75 € [-76.5; -72.8] (-19%) | -68 € [-69.3; -66.1] (-17%) | -42 € [-43.6; -40.2] (-10%) |
| Other paramedical care | -11 € [-11; -10.1] (-22%) | -16 € [-15.9; -15.3] (-32%) | -7 € [-7.1; -6.4] (-13%) | -6 € [-6; -5] (-11%) |
| General practitioners | -1 € [-1.3; -0.5] (0%) | -18 € [-18.3; -17.4] (-6%) | -12 € [-12.5; -11.7] (-4%) | -5 € [-5.3; -4.1] (-2%) |
| Midwifery | 0 € [-0.2; 0] (-19%) | 0 € [-0.3; -0.2] (-47%) | 0 € [-0.3; -0.1] (-41%) | 0 € [-0.3; 0] (-41%) |
| Dental care | -9 € [-9; -8.5] (-25%) | -7 € [-7; -6.5] (-18%) | -3 € [-3.5; -3.2] (-9%) | +0 € [0.1; 0.7] (+1%) |
| Maternity leave | +1 € [0.4; 2.4] (+72%) | 0 € [-0.8; 0.7] (-8%) | +0 € [-0.6; 1.4] (+30%) | +1 € [-0.3; 1.8] (+88%) |
| Other ambulatory care | -2 € [-2.6; -2.3] (-42%) | 0 € [-0.2; -0.1] (-3%) | +1 € [1; 1.2] (+23%) | +2 € [1.5; 1.8] (+40%) |
| Laboratory tests | +45 € [44.2; 45.8] (+22%) | +61 € [59.7; 61.5] (+31%) | +39 € [38.7; 40.3] (+20%) | +9 € [9; 10] (+5%) |
| Disability pension | +12 € [9.9; 15.2] (+12%) | +21 € [18.8; 23.9] (+19%) | +25 € [22.4; 27.7] (+25%) | +22 € [17.2; 27.5] (+23%) |
| Public hospital outpatient care | +1 € [0.3; 1.9] (+0%) | +8 € [7.2; 8.7] (+3%) | +30 € [29.7; 31.1] (+11%) | +42 € [40.6; 43.4] (+14%) |
| Sick leave | +21 € [16.4; 24.5] (+10%) | -1 € [-4.7; 3.3] (0%) | +24 € [20.4; 28.5] (+13%) | +51 € [44.2; 57.3] (+29%) |
| Hospitalization in psychiatry | +140 € [129.9; 154.6] (+86%) | +77 € [68.8; 87.9] (+47%) | +52 € [46.4; 58.6] (+34%) | +81 € [67.9; 95] (+53%) |
| Hospital at home | +84 € [78.6; 89.9] (+26%) | +230 € [225; 236.5] (+72%) | +151 € [142.5; 160.9] (+42%) | +86 € [79.3; 94.3] (+23%) |
| Phisiotherapists | -6 € [-6.5; -4.9] (-2%) | +58 € [56.6; 58.9] (+19%) | +70 € [68.8; 71.1] (+21%) | +90 € [87.9; 91.8] (+27%) |
| Drugs and medical devices out of DRG | -75 € [-85.1; -68.1] (-7%) | -77 € [-89.3; -64.3] (-7%) | +176 € [156.3; 194.3] (+14%) | +310 € [286; 334] (+24%) |
| Drugs | +43 € [35.8; 50.8] (+2%) | +76 € [67.7; 83] (+4%) | +324 € [309.7; 344.9] (+18%) | +316 € [303.2; 325.6] (+17%) |
| Medical devices and associated care | +179 € [175.3; 182.7] (+21%) | +270 € [265.4; 275.4] (+32%) | +357 € [351.3; 362.9] (+41%) | +352 € [344.8; 359.7] (+39%) |
| Transportation | +152 € [146.3; 158.1] (+19%) | +248 € [242.5; 256.2] (+33%) | +366 € [358.2; 373.2] (+47%) | +416 € [408; 424.7] (+54%) |
| Nursing care | +193 € [187.7; 197.9] (+13%) | +274 € [266; 280.3] (+18%) | +449 € [441.6; 455.6] (+27%) | +425 € [416.3; 433] (+25%) |
| Short-stay hospitalization (DRG*) | +6755 € [6633.9; 6870.5] (+56%) | +6959 € [6824.9; 7079.6] (+59%) | +6204 € [6073.8; 6318] (+51%) | +7167 € [7019.4; 7328.8] (+60%) |
| Hospitalization in rehabilitation care | +5400 € [5295.8; 5484.2] (+173%) | +4590 € [4510.6; 4666.3] (+151%) | +6849 € [6759.6; 6929.7] (+219%) | +8254 € [8111.2; 8440.3] (+264%) |

Year

*DRG: diagnosis-related group.

For each year and expenditure category, the color gradient reflects the estimated excess expenditure by patient among ACVD patients with at least one COVID-19-related hospitalization during the year. The average amount is reported (with 95% confidence interval), as well as the relative percentage compared to expected values. For example, in 2020, observed expenditures for short-stay hospitalization per patient were €6755 above those expected based on pre-pandemic years. This excess expenditure corresponded to a rise of 56% compared to expected expenditures.

**Supplementary Figure 13. Estimated observed minus expected expenditure by category, ACVD COVID-, France, 2020–2023**

| Expenditure category | 2020 (-151 M€) | 2021 (+286 M€) | 2022 (+329M€) | 2023 (+896M€) |
|---|---|---|---|---|
| General practitioners | -1.7 M€ [-1.7; -1.7] (-2%) | -3.6 M€ [-3.6; -3.5] (-3%) | -8.7 M€ [-8.7; -8.6] (-8%) | -8.5 M€ [-8.6; -8.5] (-8%) |
| Public hospital outpatient care | -12.4 M€ [-12.5; -12.4] (-12%) | -2.7 M€ [-2.7; -2.7] (-2%) | -2.9 M€ [-3; -2.9] (-3%) | -0.3 M€ [-0.4; -0.3] (0%) |
| Midwifery | -60167 € [-61719; -58261] (-19%) | -23887 € [-26156; -21607] (-7%) | -11281 € [-13446; -9435] (-3%) | -3033 € [-5787; 87] (-1%) |
| Maternity leave | +11375 € [-9040; 25174] (+1%) | +0.1 M€ [0.1; 0.1] (+6%) | +0.2 M€ [0.2; 0.2] (+10%) | +0.2 M€ [0.2; 0.3] (+12%) |
| Other paramedical care | -1.5 M€ [-1.5; -1.4] (-8%) | -0.6 M€ [-0.6; -0.6] (-3%) | -0.1 M€ [-0.2; -0.1] (-1%) | +0.5 M€ [0.5; 0.5] (+3%) |
| Disability pension | -1 M€ [-1.1; -1] (-1%) | -0.8 M€ [-0.8; -0.7] (-1%) | +1 M€ [0.9; 1.1] (+1%) | +1.4 M€ [1.3; 1.5] (+2%) |
| Other ambulatory care | -1.5 M€ [-1.5; -1.5] (-57%) | +2.1 M€ [2.1; 2.1] (+80%) | +1.3 M€ [1.3; 1.3] (+54%) | +1.5 M€ [1.5; 1.5] (+66%) |
| Dental care | -1.5 M€ [-1.5; -1.5] (-9%) | +2.5 M€ [2.5; 2.5] (+14%) | +3.1 M€ [3.1; 3.1] (+17%) | +3.8 M€ [3.7; 3.8] (+20%) |
| Laboratory tests | +13.4 M€ [13.3; 13.4] (+18%) | +24.4 M€ [24.3; 24.5] (+33%) | +15.4 M€ [15.3; 15.4] (+22%) | +4.8 M€ [4.8; 4.8] (+7%) |
| Specialists | -5.1 M€ [-5.2; -5.1] (-3%) | +4.9 M€ [4.9; 5] (+3%) | +3.9 M€ [3.8; 3.9] (+2%) | +6.6 M€ [6.5; 6.6] (+4%) |
| Phisiotherapists | -8.3 M€ [-8.3; -8.3] (-8%) | +4.4 M€ [4.4; 4.4] (+4%) | +3.8 M€ [3.8; 3.9] (+4%) | +8.5 M€ [8.5; 8.5] (+8%) |
| Nursing care | +20 M€ [19.9; 20.1] (+4%) | +13.1 M€ [13; 13.2] (+3%) | +9.7 M€ [9.5; 9.8] (+2%) | +8.8 M€ [8.7; 9] (+2%) |
| Hospital at home | +12.1 M€ [11.9; 12.2] (+12%) | +10.2 M€ [10; 10.4] (+10%) | +0.7 M€ [0.6; 0.9] (+1%) | +11.9 M€ [11.7; 12.1] (+10%) |
| Drugs and medical devices out of DRG | -1.8 M€ [-2.2; -1.3] (0%) | +21.3 M€ [20.8; 21.8] (+4%) | +22.7 M€ [22.1; 23.5] (+5%) | +13 M€ [12.1; 13.8] (+2%) |
| Sick leave | +0.2 M€ [0.2; 0.3] (+0%) | -4.2 M€ [-4.2; -4] (-2%) | +7 M€ [6.8; 7.1] (+4%) | +13.9 M€ [13.8; 14.1] (+7%) |
| Medical devices and associated care | +11.2 M€ [11.1; 11.3] (+4%) | +18.8 M€ [18.7; 18.9] (+6%) | +13.7 M€ [13.6; 13.8] (+5%) | +16.6 M€ [16.5; 16.8] (+5%) |
| Hospitalization in psychiatry | +6.6 M€ [6.4; 6.8] (+9%) | +14.3 M€ [14; 14.5] (+19%) | +20.1 M€ [19.8; 20.3] (+28%) | +18.2 M€ [17.9; 18.5] (+25%) |
| Transportation | -25.2 M€ [-25.3; -25.1] (-9%) | +22.9 M€ [22.8; 23.1] (+8%) | +34.5 M€ [34.3; 34.7] (+13%) | +62.5 M€ [62.2; 62.8] (+24%) |
| Drugs | +34.3 M€ [34; 34.6] (+5%) | +82.7 M€ [82.3; 83.2] (+13%) | +100.5 M€ [100; 101] (+16%) | +126.6 M€ [125.9; 127.2] (+19%) |
| Hospitalization in rehabilitation care | +9 M€ [8.6; 9.4] (+1%) | +39 M€ [38.4; 39.4] (+3%) | +40.5 M€ [39.9; 41.1] (+4%) | +140.6 M€ [139.9; 141.5] (+12%) |
| Short-stay hospitalization (DRG*) | -198.1 M€ [-199; -197.2] (-4%) | +36.7 M€ [34.7; 38.4] (+1%) | +62.9 M€ [61.3; 64.6] (+1%) | +465 M€ [462.4; 467.3] (+10%) |

Year

*DRG: diagnosis-related group.

For each year and expenditure category, the color gradient reflects the total estimated excess expenditure among ACVD patients without COVID-19-related hospitalization during the year. Excess expenditure is defined as the difference between the observed amount and the expected amount based on pre-pandemic trends (2015–2019) over the 2020–2023 period. The crude amount is reported (with 95% confidence interval), as well as the relative percentage compared to expected values. For example, in 2023, observed expenditures for short-stay hospitalization were €465 million above those expected based on pre-pandemic years. This excess expenditure corresponded to a rise of 10% compared to expected expenditures.

**Supplementary Figure 14. Estimated observed minus expected expenditure by category and by patient, ACVD COVID-, France, 2020–2022**

| Expenditure category | 2020 (-369 € / patient) | 2021 (+673 € / patient) | 2022 (+805€ / patient) | 2023 (+2147€ / patient) |
|---|---|---|---|---|
| General practitioners | -4 € [-4.3; -4.2] (-2%) | -8 € [-8.4; -8.3] (-3%) | -21 € [-21.2; -21.1] (-8%) | -20 € [-20.5; -20.4] (-8%) |
| Public hospital outpatient care | -30 € [-30.5; -30.2] (-12%) | -6 € [-6.5; -6.3] (-2%) | -7 € [-7.2; -7.1] (-3%) | -1 € [-0.8; -0.7] (0%) |
| Midwifery | 0 € [-0.2; -0.1] (-19%) | 0 € [-0.1; -0.1] (-7%) | 0 € [0; 0] (-3%) | 0 € [0; 0] (-1%) |
| Maternity leave | +0 € [0; 0.1] (+1%) | +0 € [0.2; 0.4] (+6%) | +0 € [0.4; 0.5] (+10%) | +1 € [0.5; 0.6] (+12%) |
| Other paramedical care | -4 € [-3.6; -3.5] (-8%) | -1 € [-1.4; -1.3] (-3%) | 0 € [-0.4; -0.3] (-1%) | +1 € [1.2; 1.3] (+3%) |
| Disability pension | -3 € [-2.6; -2.4] (-1%) | -2 € [-1.9; -1.6] (-1%) | +2 € [2.3; 2.7] (+1%) | +3 € [3.2; 3.6] (+2%) |
| Other ambulatory care | -4 € [-3.7; -3.7] (-57%) | +5 € [5; 5] (+80%) | +3 € [3.2; 3.2] (+54%) | +4 € [3.6; 3.7] (+66%) |
| Dental care | -4 € [-3.7; -3.6] (-9%) | +6 € [5.8; 5.8] (+14%) | +8 € [7.5; 7.6] (+17%) | +9 € [9; 9] (+20%) |
| Laboratory tests | +33 € [32.6; 32.7] (+18%) | +58 € [57.4; 57.7] (+33%) | +38 € [37.5; 37.7] (+22%) | +11 € [11.4; 11.5] (+7%) |
| Specialists | -13 € [-12.6; -12.4] (-3%) | +12 € [11.5; 11.8] (+3%) | +10 € [9.4; 9.6] (+2%) | +16 € [15.6; 15.9] (+4%) |
| Phisiotherapists | -20 € [-20.3; -20.2] (-8%) | +10 € [10.3; 10.5] (+4%) | +9 € [9.3; 9.5] (+4%) | +20 € [20.3; 20.5] (+8%) |
| Nursing care | +49 € [48.6; 49] (+4%) | +31 € [30.7; 31.2] (+3%) | +24 € [23.3; 24] (+2%) | +21 € [20.8; 21.5] (+2%) |
| Hospital at home | +29 € [29.1; 29.7] (+12%) | +24 € [23.6; 24.6] (+10%) | +2 € [1.4; 2.1] (+1%) | +29 € [28.1; 29.1] (+10%) |
| Drugs and medical devices out of DRG | -4 € [-5.4; -3.1] (0%) | +50 € [48.9; 51.4] (+4%) | +55 € [54; 57.4] (+5%) | +31 € [29; 33] (+2%) |
| Sick leave | +1 € [0.4; 0.8] (+0%) | -10 € [-10; -9.5] (-2%) | +17 € [16.7; 17.4] (+4%) | +33 € [33.1; 33.8] (+7%) |
| Medical devices and associated care | +27 € [27.2; 27.6] (+4%) | +44 € [44; 44.5] (+6%) | +33 € [33.2; 33.7] (+5%) | +40 € [39.6; 40.2] (+5%) |
| Hospitalization in psychiatry | +16 € [15.6; 16.7] (+9%) | +34 € [32.9; 34.1] (+19%) | +49 € [48.4; 49.7] (+28%) | +44 € [43; 44.4] (+25%) |
| Transportation | -61 € [-61.8; -61.1] (-9%) | +54 € [53.6; 54.4] (+8%) | +84 € [83.9; 85] (+13%) | +150 € [149.1; 150.5] (+24%) |
| Drugs | +84 € [83; 84.4] (+5%) | +195 € [193.9; 196.1] (+13%) | +246 € [244.5; 246.9] (+16%) | +303 € [302; 304.9] (+19%) |
| Hospitalization in rehabilitation care | +22 € [21; 23] (+1%) | +92 € [90.4; 92.7] (+3%) | +99 € [97.5; 100.4] (+4%) | +337 € [335.3; 339.2] (+12%) |
| Short-stay hospitalization (DRG*) | -483 € [-485.4; -480.9] (-4%) | +87 € [81.7; 90.4] (+1%) | +154 € [149.8; 158] (+1%) | +1115 € [1108.7; 1120.3] (+10%) |

Year

*DRG: diagnosis-related group.

For each year and expenditure category, the color gradient reflects the estimated excess expenditure by patient among ACVD patients without COVID-19-related hospitalization during the year. The average amount is reported (with 95% confidence interval), as well as the relative percentage compared to expected values. For example, in 2023, observed expenditures for short-stay hospitalization per patient were €1115 above those expected based on pre-pandemic years. This excess expenditure corresponded to a rise of 10% compared to expected expenditures.

1. Méthode [Internet]. [cited 2025 Feb 18]. Available from: https://www.assurance-maladie.ameli.fr/etudes-et-donnees/par-theme/pathologies/cartographie-assurance-maladie/methode-cartographie-pathologies-depenses-assurance-maladie

2. Rachas A, Gastaldi-Ménager C, Denis P, Barthélémy P, Constantinou P, Drouin J, et al. The Economic Burden of Disease in France From the National Health Insurance Perspective. Med Care. 2022;60:655–64.

3. Yoshida K, Hernández-Díaz S, Solomon DH, Jackson JW, Gagne JJ, Glynn RJ, et al. Matching Weights to Simultaneously Compare Three Treatment Groups: Comparison to Three-way Matching. Epidemiology. 2017;28:387.

4. Rey G, Jougla E, Fouillet A, Hémon D. Ecological association between a deprivation index and mortality in France over the period 1997 – 2001: variations with spatial scale, degree of urbanicity, age, gender and cause of death. BMC Public Health. 2009;9:33.

5. Constantinou P, Tuppin P, Fagot-Campagna A, Gastaldi-Ménager C, Schellevis FG, Pelletier-Fleury N. Two morbidity indices developed in a nationwide population permitted performant outcome-specific severity adjustment. J Clin Epidemiol. 2018;103:60–70.
